# Genomic Surveillance of Third-Generation Cephalosporin-Resistant *Klebsiella pneumoniae* in Tunisian AMR Surveillance System Hospitals

**DOI:** 10.64898/2026.04.08.26350452

**Authors:** Dana Itani, Hanen Smaoui, Lamia Thabet, Meriam Zribi, Sarra Dhraief, Lamia Kanzari, Khaoula Meftah, Wafaa Achour, Dave J Baker, Cara-Jane Moss, Laura T. Philips, Ebenezer Foster-Nyarko, Ilhem Boutiba – Ben Boubaker, Kathryn E. Holt

## Abstract

Third-generation cephalosporin (3GC) resistant *Klebsiella pneumoniae* are an increasing public health threat in Tunisia, yet there is limited data on the circulating lineages and antimicrobial resistance (AMR) determinants underlying this threat. Here, we employed whole-genome sequencing (WGS) in the Tunisian AMR surveillance system (TARSS) to characterize the 3GC resistance mechanisms, population structure, virulence, and transmission across three participating sentinel hospitals in Tunis and Ben Arous.

We sequenced a balanced sample of stored 3GCR (3GC-resistant) isolates from blood and urine collected between 2018 and 2022. Of 322 sequenced isolates, 286 (89%) were confirmed as *K. pneumoniae*, representing 28.5% of all stored 3GCR isolates. The population structure was diverse (68 sublineages) and distinct between hospitals, although several globally distributed sublineages were detected across sites (SL383, SL101, SL307, SL15). Extended-spectrum ß - lactamases (ESBL) genes were detected in 77% of genomes, with *bla*_CTX-M-15_ (65.4%) and *bla*_CTX-M-14_ (8%) dominant at all sites and across diverse sublineages. AmpC genes occurred in 9%, and carbapenemase in 19.6% (*bla*_OXA-48_, 14.7%; *bla*_NDM-5_, 4.5%; *bla*_NDM-1_, 3.8%), with carbapenemases mainly observed amongst SL147 and SL383 at Hospital B (41.7%). Despite sequencing less than a third of the unique 3GCR infections in each hospital, we identified 24 probable nosocomial transmission clusters involving 64 isolates. Each cluster was restricted to a single hospital, although many were detected across multiple wards in the same hospital. The acquired virulence-associated locus (ICE*Kp*) encoding yersiniabactin was common (48.6%). Hypervirulence-associated markers (encoding aerobactin, salmochelin, and/or hypermucoidy) were rare (8.7%) but increasing over time. These were mostly found in sublineages in which convergence of ESBL and hypervirulence has been reported in other settings (including SL147, SL101 and SL383), suggesting international dissemination of convergent strains.

These findings show sustained ward-level nosocomial transmission of 3GCR *K. pneumoniae* lineages and site-specific differences in ESBL and carbapenemase burdens, which call for targeted infection prevention and control and for future routine integration of WGS into TARSS.

## Introduction

Bacterial antimicrobial resistance (AMR) poses a global threat to human health. In 2019, it is estimated AMR was directly responsible for 1.27 million global deaths and contributed to 4.9 million deaths, with the heaviest burden in low-and middle-income countries (LMICs) (1). Economically, the World Bank projects that AMR could result in an additional $US 1 trillion in healthcare costs by 2050 if left unaddressed (2). One pathogen of particular concern is *Klebsiella pneumoniae*, a Gram-negative bacterium that causes community- and healthcare-associated infections and is responsible for an estimated 100,000 AMR-related global deaths annually (1). Its capacity to rapidly acquire AMR genes, coupled with its association with high mortality in LMICs, has led the World Health Organization (WHO) to designate *Klebsiella pneumoniae* as a critical priority pathogen, specifically *Klebsiella pneumoniae* resistant to third-generation cephalosporins (3GC) or carbapenems (3). *K. pneumoniae* has traditionally been recognized as posing two distinct clinical threats, associated with distinct clonal lineages or multi-locus sequence types (STs): high-risk multidrug-resistant sublineages (e.g., ST147, ST307) causing healthcare-associated infections, and hypervirulent sublineages (e.g., ST23, ST86) causing community-acquired invasive infections that are typically drug-susceptible (4–7). Recent genomic studies have documented the emergence of convergent “dual threat” strains that combine both hypervirulence and multidrug resistance traits, posing significant challenges to infection control and treatment (8,9).

In 2015, the World Health Assembly endorsed the Global Action Plan (GAP) on AMR and urged countries to develop and implement national action plans (NAPs) (10), including a core objective to strengthen surveillance. While phenotypic antimicrobial susceptibility testing (AST) is the cornerstone of routine surveillance, it is insufficient to identify clonal diversity, transmission dynamics, and hypervirulent or convergent clones. Recently, the WHO recommended integrating genomic surveillance into national AMR surveillance, as it can enhance the detection, characterization, and tracking of resistant pathogen variants (11). While progress on integrating whole-genome sequencing (WGS) into AMR surveillance has been made in high-income countries, WGS-enhanced surveillance remains limited in LMICs (12–14).

Tunisia, a North African country classified as lower-middle income by the World Bank, in 2019 launched its AMR NAP and established the Tunisian AMR Surveillance System (TARSS) (15). TARSS is founded on the WHO Global Antimicrobial Resistance and Use Surveillance System (GLASS) model (16,17), collecting and analyzing routine microbiology and epidemiological data from 11 sentinel sites across the country. We recently reported AMR prevalence data for four key pathogens (*K. pneumoniae*, *Pseudomonas aeruginosa*, *Acinetobacter* spp., and *Escherichia coli*) estimated from five TARSS sites from 2014 to 2022. *K. pneumoniae* was the most common agent of bloodstream infections across all sites (median 242 per year) and showed high and increasing prevalence of resistance to 3GC antibiotics cefotaxime and ceftazidime (reaching 69% and 69.5%, respectively, in 2022). 3GC-resistant (3GCR) *K. pneumoniae* were most common amongst intensive care patients and infants and were responsible for the majority of resistance to other antibiotics (15.3% imipenem resistant, 34.3% amikacin resistant, and 70.6% gentamicin resistant; vs 0.3%, 2.8%, and 5.8%, respectively, amongst 3GC-susceptible isolates) (18).

In order to understand the *K. pneumoniae* pathogen population underlying these AMR trends in Tunisia, we sequenced 286 3GCR *K. pneumoniae* isolates collected through TARSS from three tertiary hospitals between 2018 and 2022. Our objectives were to characterize the genomic landscape of 3GC resistance, identify persistent and convergent clones underlying 3GCR infections, characterize their genotypic resistance and virulence profiles, and explore the burden and topology of nosocomial transmission and its relative contribution to the burden of healthcare-associated infection (HAI). Analyses of carbapenem resistance were also undertaken. To our knowledge, this is the first genomic epidemiological study of *K. pneumoniae* in Tunisian hospitals and demonstrates the potential for integration of genomics into national AMR surveillance to yield useful insights in this setting.

## Methods

### Ethical considerations

The study was approved by the Ethics Committee of Charles Nicolle Hospital in Tunis (Authorization No. FWA00032748 and IORG0011243) and received authorization from the Ministry of Health in Tunisia (Authorization No. 0000021-080000-20-2023), in addition to approvals from the London School of Hygiene & Tropical Medicine (LSHTM) Observational/Interventions Research Ethics Committee (Ref: 26901). A waiver of individual participant consent was granted due to the retrospective anonymized design and the use of isolates collected as part of routine national surveillance. All patient data included in the study were anonymized by National Reference Laboratory of AMR (NRL), prior to sharing with any research staff who would not normally have access to patient records. Sequence metadata deposited in public archives included only contextual metadata on the specimen type (blood or urine), year, country, and specimen collection date, with no information on hospital, admission date, gender, or age, to limit the chances of re-identification of individuals. Shipment of *K. pneumoniae* isolates to LSHTM complied with Tunisia’s national regulations governing biosafety and sharing of genetic resources, and the Nagoya Protocol. Additionally, the benefits from this research (enhanced AMR surveillance capacity, training workshop, and result dissemination) were shared with participating hospitals and the NRL.

### Study design and setting

This study combined prospectively collected surveillance data (as previously reported by Itani *et al*. (18) with retrospective genomic characterization of a subset of stored 3GCR *K. pneumoniae* surveillance isolates collected from three TARSS sites between 2018 and 2022. The study reporting adheres to STROBE (Strengthening the Reporting of Observational studies in Epidemiology) and extension STROME-ID (Strengthening the Reporting of Molecular Epidemiology for Infectious Diseases) standards (19).

TARSS monitors WHO GLASS priority pathogens including *K. pneumoniae* and other bacteria of local public health importance. Clinical specimens (blood, urine, stool, wound, respiratory, cerebrospinal fluid, and genital tract) are processed across 11 sentinel sites in Tunisia. Culture and antimicrobial susceptibility testing (AST) data are shared electronically with the NRL. The laboratories utilize two information systems: Santé Lab, Laboratory Information System (LIS) and SIRscan (automated AST reader), linked nationally after the establishment of the TARSS in 2019. To ensure laboratories provide quality-assured results, all sites participate in an external quality assessment scheme coordinated by the NRL and the National Coordinating Centre.

Three tertiary-care TARSS hospitals were selected for genomic characterization of 3GCR *K. pneumoniae* isolates based on their diverse clinical services and patient populations, high numbers of 3GCR *K. pneumoniae*, and availability of archived isolates. Hospital A (332 beds) specializes in pediatric and neonatal care in Tunis. Hospital B (930 beds) is a national referral center serving all age groups in Tunis, the capital city. Hospital C (164 beds) is a national referral center for trauma and burns in Ben Arous City, south of Tunis. **Table 1** summarizes hospital characteristics; isolate distribution and sampling workflow are shown in **Fig 1**.

**Figure 1.**
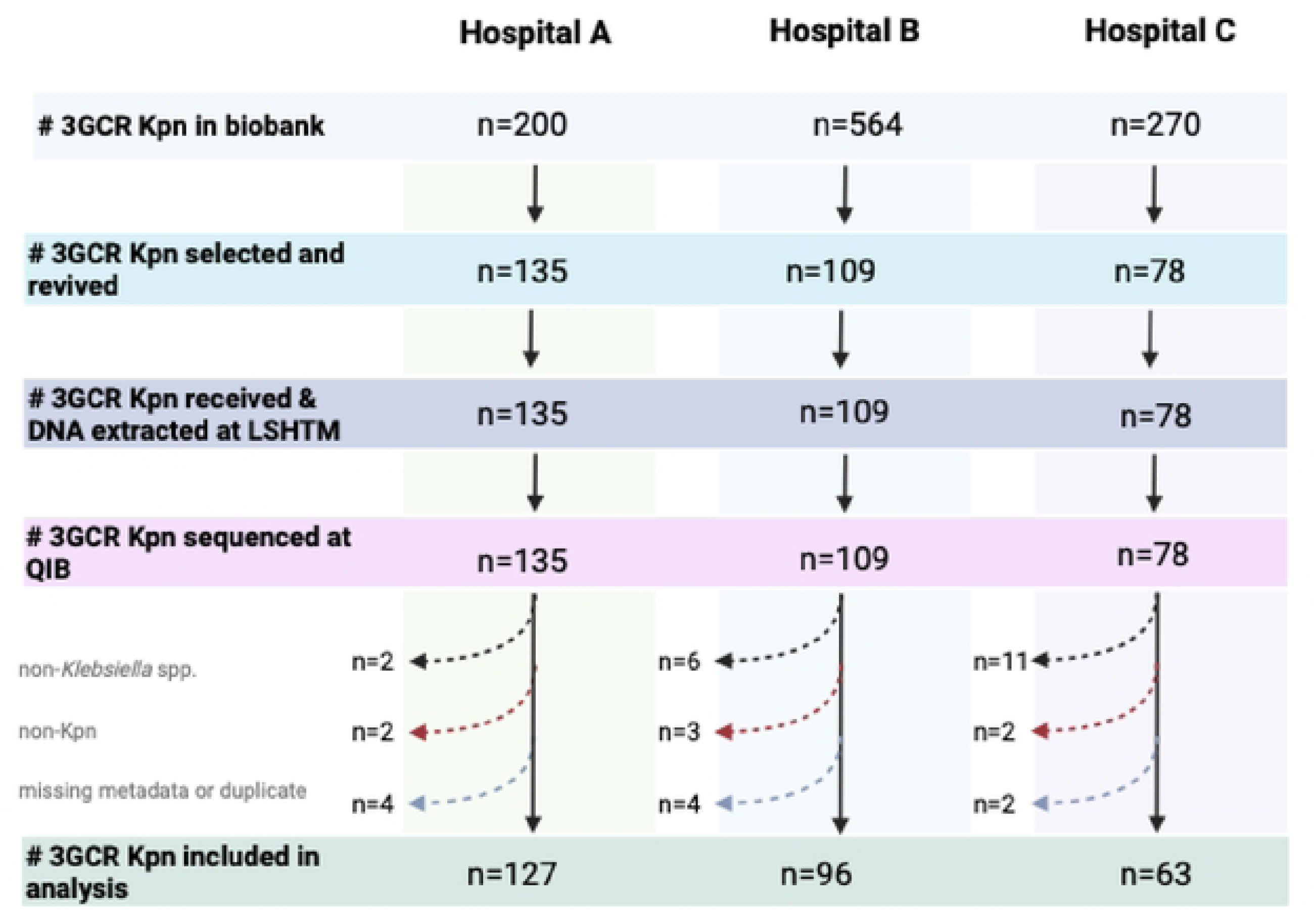
Study sites and sample selection workflow. Flowchart showing the collection, laboratory processing, sequencing, and final inclusion of third-generation ccphalosporin-rcsistant *Klebsiella pneumoniae* clinical isolates from each hospital. **Abbreviations:** LSHTM=London School of Hygiene and Tropical Medicine; QIB= Quadram Institute Bioscience; Kpn= *Klebsiella pneumoniae*

**Table 1.** Characteristics of hospitals and overview of the WGS sampling frame of3GCR *K. pneunwniae* isolates, Tunisia, 2018-2022. Stored isolates refer to the total number of third-generation cephalosporin-resistant (3GCR) *K pneumoniae* isolates from blood and urine that were archived in hospital microbiology laboratory biobanks between 2018 and 2022. After record linkage to TARSS surveillance database and application of WHO de-duplication criteria (one isolate per patient, specimen type, and year), n=1,005 isolates constituted eligible sampling frame.

| Panel A- Hospital characteristics |  |  |  |
| --- | --- | --- | --- |
| Characteristics | Hospital A | Hospital B | Hospital C |
| City | Tunis | Tunis | Ben Arous |
| Specialisation | Paediatrics and paediatrics surgery | Medical and surgical specialties | Trauma and burn care |
| Bed capacity | 332 | 930 | 164 |
| Admissions (2023) | 19,608 | 27,950 | 8,749 |
| Patient population | Children, <15 years | All ages | Primarily adults, ≥18 years |
| Panel B- Sequenced 3GCR *K. pneumoniae* (n) |  |  |  |
| 2018 | 36 | 0 | 17 |
| 2019 | 25 | 12 | 19 |
| 2020 | 18 | 27 | 10 |
| 2021 | 33 | 36 | 10 |
| 2022 | 15 | 21 | 7 |
| Panel C- Sequenced isolates by specimen type (n) |  |  |  |
| Blood | 58 | 34 | 36 |
| Urine | 69 | 62 | 27 |
| Panel D- Sequencing coverage of the eligible biobank sampling frame |  |  |  |
| Hospital | Sequenced isolates (n) | Eligible isolates in sampling frame (n) | Coverage (%) |
| Hospital A | 127 | 167 | 76 |
| Hospital B | 96 | 562 | 17.1 |
| Hospital C | 63 | 276 | 22.8 |
| Total | 286 | 1005 | 28.5 |

### Specimen collection, bacterial identification and antimicrobial susceptibility testing

TARSS collects routine diagnostic blood and urine specimens from inpatients and outpatients when suspected of infection. Bacterial identification and AST were performed locally at hospital microbiology laboratories following TARSS protocols. Species identification was conducted using colony morphology, Gram staining, and manual biochemical testing, including API20E strips (bioMérieux). AST was performed using the disk diffusion method, and disk zones were interpreted according to European Committee on Antimicrobial Susceptibility Testing (EUCAST) clinical breakpoints (version per year of use). 3GCR was defined as resistance to cefotaxime and/or ceftazidime.

Isolates confirmed as *K. pneumoniae* were subcultured and stored at -80°C in the respective hospital laboratories for surveillance and research purposes. For this study, selected non-duplicate 3GCR *K. pneumoniae* isolates (see **Isolate selection for WGS**) collected between 2018 and 2022 were revived at hospitals, and those that survived the archive were shipped to the NRL.

We used for our primary phenotypic summaries the hospital laboratories AST interpretations (S/I/R) that were made using the latest EUCAST version in use in the corresponding year. For a subset of isolates (86.4% of total sequenced isolates), disk diffusion zone diameters were also available and were reinterpreted using the AMR R package v3 using EUCAST 2024 breakpoints (20,21). The availability of zone diameters was lower in 2022 (46.5%), reflecting missing diameters reported to TARSS by Hospital B. These reinterpretations were used exclusively for UpSet analysis. We treated susceptible “S”, susceptible increased exposure “I”, and resistant “R” separately and did not combine “I” with resistant “R”, as per EUCAST guidance (22). EUCAST expected resistance and expert rules were applied where relevant. The antibiotics analyzed in this study were the 3GCs, cefotaxime and ceftazidime, and carbapenems, ertapenem and imipenem, for which AST results were available for all samples. The proportion resistant to each antibiotic was calculated by year and specimen type, and linear trends were assessed using the Cochran–Armitage trend test.

Demographic and clinical metadata, including patient age and gender, ward type, and infection type (healthcare-associated infection [HAI] vs community-acquired infections [CAI]), were extracted from Santé Lab. Samples were stratified into age groups (0–28 days, 29–365 days; 1–4 years, 5–9 years, 10–14 years, 15–19 years, 20–24 years, 25–59 years, 60–99 years, >100 years) as per Diaz *et al* (23). CAI were defined by the isolation of the pathogen from an outpatient or on days 0, 1, and 2 of current admission as an inpatient. HAI was defined as an infection in a patient that occurred after being hospitalized for >2 days (24).

### Isolate selection for WGS

Across the three participating hospitals, the laboratory biobank contained 1,034 vials identified as 3GCR *K. pneumoniae* isolated from blood or urine samples between 2018 and 2022. After record linkage to the TARSS database and application of WHO de-duplication criteria (unique sample per patient and specimen type in the same year (25), 1,005 isolates constituted the sampling frame. The difference (n=29) reflects the biobank entries that were unmatched to TARSS or duplicates. Sufficient resources were available to sequence 300–350 isolates (approximately one-third of the total stored isolates available). To enable year-on-year comparisons of the prevalence of distinct 3GCR *K. pneumoniae* clones, we aimed to balance equal sample size per year (2018–2022), per hospital (Hospital A, Hospital B, Hospital C), and per specimen type (blood vs urine). However, no isolates were retrieved from Hospital B in 2018; no blood isolates were retrieved from Hospital A in 2020; and at Hospital C, only four urine isolates were recoverable by 2021; while in 2022, only a single blood isolate could be retrieved. Given these constraints, we selected and revived 322 3GCR *K. pneumoniae* isolates for WGS analysis, totalling 43–79 genomes per year and providing sufficient temporal and site-level coverage for comparative analysis (**Table 1**). Isolate-level linkage between biobank inventory and TARSS records was not available for stratified analysis, therefore the comparisons between TARSS and WGS distributions are descriptive and should not be interpreted as estimates of surveillance coverage or representativeness.

### DNA extraction and whole genome sequencing

The surveillance data from TARSS used in this study were accessed on 17 January 2023 for research purposes following local ethics approval. The isolates and associated sequencing metadata were subsequently accessed on 12 January 2024. At the time of the study, WGS capacity was not yet available at the NRL, therefore the selected 3GCR *K. pneumoniae* isolates were shipped to LSHTM, UK, for genomic analysis. At LSHTM, isolates were cultured on MacConkey agar (Fisher Scientific, UK); a single colony per plate was then subcultured, and genomic DNA was extracted following the 96-well lysate protocol by Foster-Nyarko *et al.* (26). DNA was assessed for purity (A260/A280 and A260/A230) using the NanoDrop 2000 Spectrophotometer (Fisher Scientific, Loughborough, UK) and concentration was quantified using the Qubit high-sensitivity DNA assay (Invitrogen, MA, USA). DNA extracts were stored at -20°C and then transferred to the Quadram Institute Bioscience (UK) for DNA library preparation and sequencing.

For library preparation, DNA was quantified using Qubit and normalized to a concentration of 5 ng/μl. Libraries were prepared by tagmentation with bead-linked transposomes and tagmentation buffer, and indexed with unique dual indices following Baker *et al.* (27). Library fragment size and concentration were assessed using Agilent Tapestation 4200, and libraries were normalized to approximately 0.5 nM prior to pooling. Pooled libraries were sequenced on an Illumina NextSeq 2000 platform with P3 X-Leap flow cell (300 cycles) with a 1% PhiX control spike, to generate 2x 150 bp paired-end reads.

### Bioinformatics analysis

Raw reads (FASTQ files) were analyzed using Pathogenwatch (accessed in May 2024), an open-access platform for automated bacterial genomic epidemiology analysis (28). Pathogenwatch was used for *de novo* genome assembly with SPAdes v3.15 and assembly quality checks with QUAST v5.0. Genomes were considered to be of suitable quality if they met all of the following criteria: N50 ≥15,000; genome length between 4.6 megabases and 6.4 megabases; total number of contigs ≤500; percentage of guanine (G) and cytosine (C) between 56.35–57.98 (29). Species identification was confirmed by Pathogenwatch’s Speciator module, which applies Mash to compare each assembly against a curated database of reference sequences.

We used the 629-locus *Klebsiella* core-genome multilocus sequence typing (cgMLST) scheme, curated in the BIGSdb-Pasteur database and implemented in Pathogenwatch, to determine genetic relatedness among *K. pneumoniae* sequences (30). We reported sublineage (SL), clonal group (CG), and 7-locus MLST sequence type (ST) (31). Pairwise core-gene SNP distances were obtained from Pathogenwatch, which profiles each *K. pneumoniae* genome against a species-specific core gene library (1,972 genes; 2,172,367 bp) and aggregates single-nucleotide substitutions across matched core loci to derive a core-gene SNP distance matrix (28). We did not apply any additional recombination masking. Neighbor-joining trees were generated within Pathogenwatch from the core-gene SNP distance matrix. Resulting Newick-format tree files were exported and visualized in R using ggtree v3.6.2 and ggtreeExtra v1.8.1 (32).

Genotyping for AMR and virulence determinants was conducted within Pathogenwatch using Kleborate v3.1.0 to identify acquired AMR genes, AMR-associated chromosomal mutations and virulence loci (33), and Kaptive v3.0.0b6 for capsule (K) and lipopolysaccharide (O) serotype prediction (34).

### Clonal diversity

To quantify clonal diversity, we stratified the data by hospital and specimen type. For each stratum, we tabulated SL counts and calculated the Simpson’s diversity index, D, using diversity function in the vegan v2.6-4 R package (35). The value of D can range from 0 (monoclonal population) to 1 (polyclonal highly diverse abundances). Uncertainty in D was estimated by non-parametric bootstrap resampling of isolates within each stratum (5,000 resamples) to obtain 95% confidence intervals (CI) from 2.5^th^ and 97.5^th^ percentiles. Strata (i.e. hospital-specimen type combinations) with fewer than 2 isolates were not assessed for diversity. To test whether any SL was overrepresented in urine versus blood specimen types, we used two-sided exact binomial tests; p-values were Bonferroni-adjusted across SLs.

We evaluated concordance among typing schemes (SL, CG, and ST) in two ways: (i) heterogeneity within SL, for each SL we summarized its composition in terms of ST and CG by calculating the number of distinct STs or CGs observed within that SL, and proportion of isolates belonging to the dominant ST or CG in that SL. (ii) We used the Adjusted Rand Index (ARI) to measure the agreement between SL and ST using *adjustedRandIndex* function in *mclust* v6.0.1.

### Mechanisms of 3GCR

To identify 3GCR mechanisms from WGS data, we first considered AMR genotype data reported by Kleborate (33), which includes the detection of extended-spectrum β-lactamases (ESBL), AmpC ß-lactamase, carbapenemase genes, ESBL-associated mutations in the intrinsic (core gene) ß-lactamase *bla*_SHV_ (36), and resistance-associated disruption of outer membrane porins (OmpK35, OmpK36) (33). Detection of *bla*_TEM-1_ was classified as non-ESBL (37); while for other *bla*_TEM_ alleles, ESBL status was determined by the presence of established amino acid mutations (E104K, R164S/H, G238S, E240K, A237T) (38), based on allele definitions curated in the Beta-Lactamase DataBase (BLDB) (39).

For isolates without any ESBL, AmpC, or carbapenemase detected, we reviewed WGS assembly quality and AST data, designating disk zones within 1–2 mm of the breakpoints for cefotaxime and ceftazidime as dubious results. In order to detect the presence of genes that may have been missed due to genome assembly issues, we also screened the raw unassembled reads directly for ß-lactamases using a local copy of the ResFinder gene database (downloaded October 2025). Paired-end reads were aligned to the gene database using Bowtie2 v2.5.4 (in end-to-end mode with --very-sensitive preset). SAM/BAM processing was performed with SAMtools v1.22.1. For each target gene, we calculated percent coverage and nucleotide identity using Python v3.13.7. Genes were considered present if they had ≥80% coverage and ≥90% identity in this reads-based analysis, and/were reported as present by Kleborate assembly-based analysis.

Based on the combined results of Kleborate and reads-based analyses, we classified the 3GCR mechanism for each isolate as follows: *bla*_CTX-M_-only; *bla*_CTX-M_ + *bla*_SHV_ mutation ± *bla*_TEM_ mutation; porin mutation only; *bla*_AmpC_ beta-lactamase; carbapenemase + porin mutation; carbapenemase + porin mutation + *bla*_AmpC_; carbapenemase only; none. The distribution of AST phenotypes vs combinations of 3GCR mechanisms were visualized using the AMRgen R package v0.0.1.9000 (40).

For each gene or gene family, we tested prevalence across hospitals/specimens using ^2^ tests or Fisher’s exact tests when counts were <5. For significant genes following adjustment, pairwise hospital and specimen comparisons were performed using Fisher’s exact test; all p-values were Holm-adjusted. Differences in inhibition zone diameters for each 3GCR drug between isolates harboring resistance determinants and those lacking were assessed using the Wilcoxon rank sum test in stats v 4.2.2. The positive predictive value (PPV) for genotype-phenotype concordance was obtained from *AMRgen* output.

### Virulence gene profiling

Following Lam *et al.* (33), we defined convergence as a virulence score of at least 3 (which corresponds to the presence of the aerobactin synthesis locus (*iuc*) (4), amongst our 3GCR isolates. The yearly proportion of *iuc*-positive isolates among 3GCR isolates was calculated, and trends were evaluated using a two-sided Cochran–Armitage trend test. We additionally calculated the proportion of *iuc*-positive isolates by specimen type, hospital, and ward group (ICU vs non-ICU), infection type (HAI vs CAI), and *K. pneumoniae* SL. Group comparisons (ICU vs non-ICU; HAI vs CAI) were performed using Fisher’s exact test (two-sided).

### Selection of genetic and temporal thresholds for putative transmission

We used the Pathogenwatch pairwise single-nucleotide polymorphism (SNP) distance matrix (described above) as the measure of pairwise genetic distance between isolates. Since this approach is assembly-based, reference-guided, and does not explicitly mask recombination, we calibrated thresholds empirically rather than assuming a fixed mutation rate on all isolates. We evaluated a grid of genetic distance thresholds (SNP ≤ {10, 12, 15, 18, 20, 25, 30, 35, 40}) and temporal distance thresholds (days ≤ {28, 56, 84}) based on isolate collection date of all sequenced isolates. For each SNP-window pair, we constructed an undirected proximity graph in which isolates *i* and *j* were linked when both criteria were met.

For each pair of SNP and temporal thresholds, we computed:

1. Proportion of isolates in clusters:

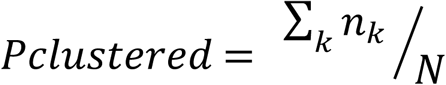 Where *n_k_* is the size of cluster *k* (connected component nk≥2) and *N* is the total number of isolates analyzed.
2. Proportion attributable to transmission

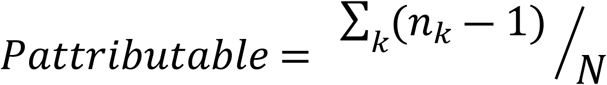
3. Number of clusters and median cluster size
4. Cluster duration (the time between the first and last specimen collection dates for all isolates in the cluster).
5. Specificity by ward (mono-ward share) defined as the proportion of clusters whose isolates all came from the same ward.
6. Pair-level ward concordance: among all edges (eligible isolate pairs) at a given cut-off, we calculated the fraction originating from the same ward within the same hospital.

To assess whether the observed within-ward clustering was greater than expected by chance, we performed 500 permutations of ward labels within the same hospital, year, and month, recalculating the fraction of same-ward pairs each time. We summarized the null by its mean and upper quantiles (95^th^ and 99^th^). Observed concordance exceeding 95–99% null range was interpreted as non-random within-ward clustering.

We assessed stability from three perspectives: (i) Temporal window stability. For each SNP threshold, we compared metrics at 56 vs 84 days (i.e., 8 vs 12 weeks) and recorded the change. The shortest window whose metrics were indistinguishable from the longer period was preferred. (ii) Cluster membership stability. At 8 weeks (56 days), we calculated per-isolate agreement in cluster membership between adjacent SNP thresholds (15 vs 18, 18 vs 20, etc.). Agreement was defined as the fraction of overlapping isolates assigned to the same component at both thresholds; values near 1 indicated stability, whilst values near 0 indicated extensive reconfiguration. (iii) Lineage composition stability, using SL. We compared the set of SLs represented in the cluster across thresholds. For each SNP threshold, coverage (S) was calculated relative to the broadest threshold (SNP≤40): 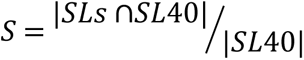, where SL_40_ is the set of SL at the broadest threshold (SNP≤40).

Adjacent Jaccard similarity 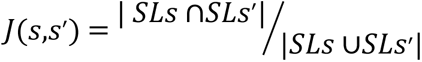 for consecutive thresholds *s* and *s*’. High coverage and high adjacent Jaccard indicate stabilization of strain composition.

In order to select thresholds of temporal window and genetic distances to define putative transmission links, we pre-specified rules prioritizing stability and specificity. For selecting the temporal window, we chose the shortest window for which extending the window produced minimal changes in key metrics (percentage clustered, number of clusters, median span). Second, we fixed the temporal window and selected the SNP threshold taking into consideration the following: (i) to avoid over-merging, we selected the threshold immediately before over-merging point; (ii) lineage specificity, aiming for high adjacent Jaccard similarity (≥0.9) between thresholds and high coverage (≥95%) of final SL set; (iii) evidence of ward localisation, based on the permutation test, aiming for observed same-ward fraction exceeding the 95% percentile of the permuted null. By applying these rules to the observed set, we selected a primary definition of putative transmission as ≤20 SNPs and ≤56 days. For our cluster analysis, we excluded CAI isolates and retained only HAI cases (60% of the total sequenced, representing 40% of all recorded de-duplicated HAI 3GCR *K. pneumoniae* isolates).

### Statistical modelling of factors associated with clustering

Multivariable logistic regression was used to investigate the association of biological and hospital factors and cluster membership. The dependent variable was defined as belonging to a cluster under our primary definition of putative transmission (clustered =1, unclustered = 0). Only HAI cases were included. We chose predictors based on biological plausibility and hospital epidemiology: presence of ESBL or carbapenemase, presence of yersiniabactin or aerobactin loci, sex (male vs female), hospital, ward (medical, surgical, ICU and other [combining outpatient and emergency wards]), and specimen type (blood vs urine). We did not include year because sampling was designed to be distributed across years and to preserve degrees of freedom. We fitted the predictors using a bias-reduced generalized linear model brglm v0.9.2 (type=AS_mean). We set the following reference categories: absence of ESBL and carbapenemase genes, absence of virulence genes (aerobactin and yersiniabactin), male sex, blood specimen, medical ward, and Hospital B as the reference hospital. We report odds ratios with 95% CI and p-values.

### Global SL147 phylogeny and annotation

In October 2025, we downloaded all SL147 *K. pneumoniae* assemblies available in Pathogenwatch (n=2,591). Missing SL147 paired-end reads from a European cross-border surveillance study (n = 87) (41) were retrieved from the European Nucleotide Archive (ENA) (BioProject accessions: PRJEB35890, PRJEB60743, PRJEB74083, PRJEB75178, PRJEB89771, PRJEB89810, PRJNA1076808, PRJNA1106484, PRJNA1143178, PRJNA288601, PRJNA648389, PRJNA657553, PRJNA903550, PRJEB89896, PRJEB76821, and PRJNA1268013). We then assembled the retrieved genomes using SPAdes v4.2.0 (default settings), yielding a total of 2,678 assemblies.

For the global phylogeny tree, we used an alignment-free k-mer-based approach. Pairwise genome distances were computed with Mash v2.3 (default k-mer size k=21; sketch size s=10,000) via Mashtree v1.2.0 (Bioconda) (42). The Mash distance matrix was converted to PHYLIP format, and a neighbor-joining tree was inferred using Quicktree v2.5 (43). Trees were handled as unrooted and visualised in R using ggtree v3.6.2 (44).

We defined co-occurrence as the simultaneous presence of aerobactin (*iuc*) and *bla*_OXA-48_. To summarise how dispersed these co-occurrences were across the global SL147 phylogeny, we grouped tips by phylogenetic proximity and counted the number of distinct clades containing ≥1 both-positive tip (denoted K). Clades were obtained by average-linkage hierarchical clustering using the hclust function in R stats v4.2.*2*. The dendrogram was cut at a fixed height equal to 2% of the tree diameter (h = 0.02x maximum patristic distance). This threshold aggregated tips at a deep sublineage scale and avoided inflating of K by splitting near-duplicate shallow clusters. We repeated this analysis to count the number of distinct clusters containing ≥1 Tunisian *iuc*-positive tip. The global phylogeny tree and its associated metadata are available in Microreact (https://microreact.org/project/pHeiJxFXa2qBQMGo1SJAso-sl147-global-tree-2026).

### Assembly-graph localization of plasmid markers

In order to check the location of resistance and virulence genes, we used SPAdes assemblies for all SL147 isolates as described above. A reference sequence for *bla*_OXA-48_ (GenBank AY236073.2) was retrieved from the National Center for Biotechnology Information (NCBI), as well as the aerobactin locus (*iucA,B,C,D*, and *iutA*) from the Virulence Factor Database, an IncHI1B_pNDM_MAR plasmid replicon marker sequence from PlasmidFinder, and the 59-mer backbone marker: (ATTCCGAACATAAATGCAATGATGAGCAGTAAGAGCACGCCCATTTGCAGCGCCGG AAG) reported by Linkevicius *et al.* as diagnostic of the hybrid *bla*_OXA-48_/*iuc* plasmid (41). Using the assembly graph viewer Bandage v0.8.1, we queried our assemblies using nucleotide BLAST with a ≥90% identity and E-value ≤1×10^-20^. We considered the mosaic plasmid present only when all four markers (*bla*_OXA-48_, aerobactin locus, IncHI1B_pNDM_MAR, and the 59-mer backbone marker) were present within a single path through the assembly graph as defined by Linkevicius *et al.* (45).

### Data Availability

The WGS sequences generated in this study were deposited in the European Nucleotide Archive (ENA) under BioProject PRJEB98550. Individual sample accessions for data used in this project are given in the **Supplementary Excel File**. The analysis code to generate results and figures is available on GitHub (DOI: 10.5281/ZENODO.19055814) (46).

## Results

### Study population and WGS sampling frame

At the three hospitals studied, the proportion resistant to 3GCs (ceftazidime or cefotaxime) was consistently higher amongst *K. pneumoniae* blood isolates compared with urine isolates, across the full surveillance period 2014–2022 (see **Fig 2**). This is consistent with previously reported analysis of an extended TARSS dataset including two additional hospitals (18). A total of 322 3GCR *K. pneumoniae* isolates were revived and subjected to WGS. Of these, 286 (89%) were confirmed from genome data as *K. pneumoniae* genomes and passed quality control filters, and were included in the analysis. The excluded sequences were mostly non-*Klebsiella* species (19/322, 5.9%) or *Klebsiella* species other than *K. pneumoniae* (7/322, 2%). The remainder (10/322, 3%) had no clinical metadata or were identified from the metadata as duplicates.

**Figure 2.**
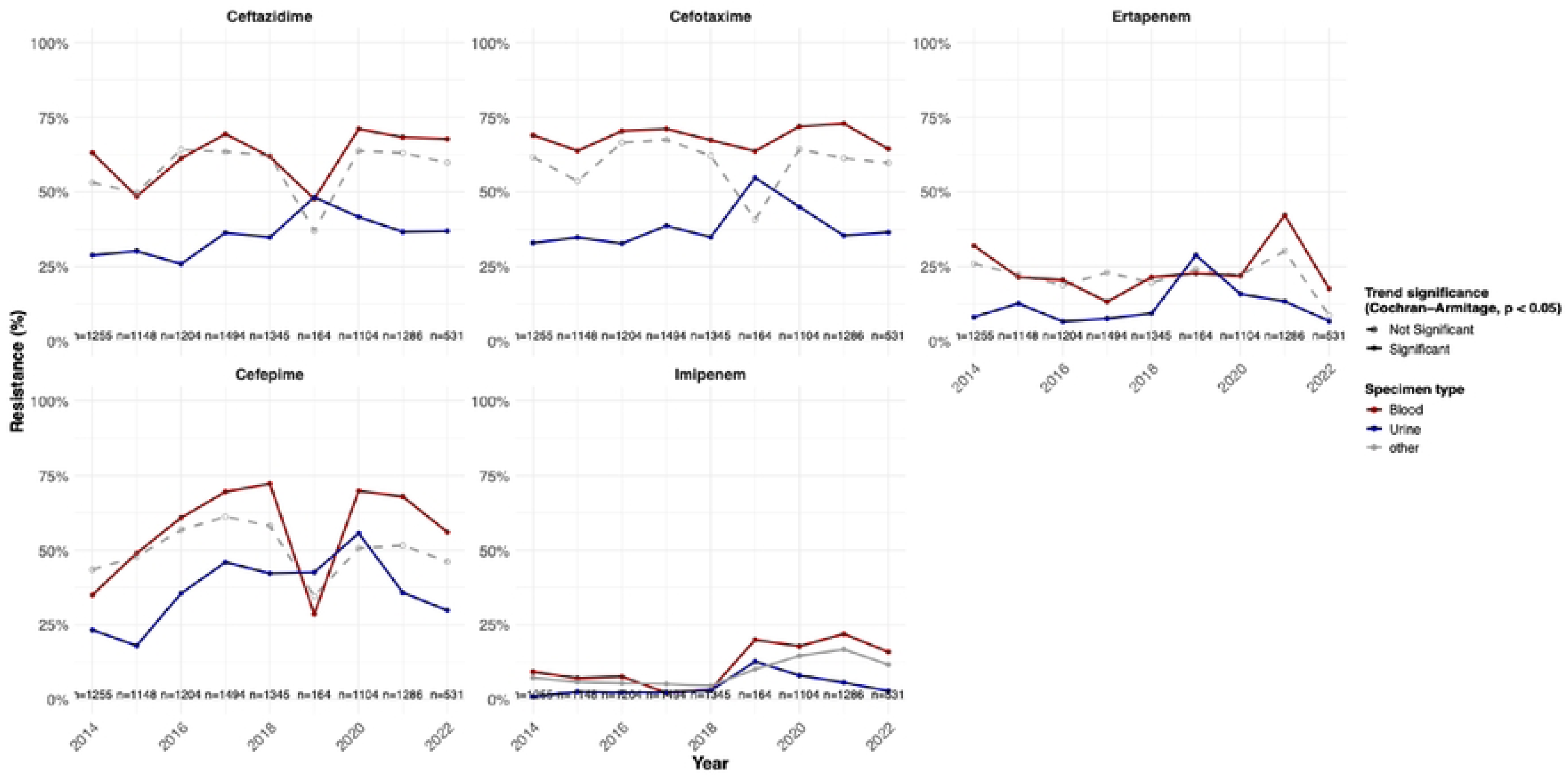
Trends in *K. pneu111oniae* resistance by specimen type across three selected TARSS hospitals, 2014-2022. The shaded area indicates the period from which stored isolates were later selected for WGS (2018–2022). Solid lines indicate statistically significant trend (Cochran-Annitage test, p<0.05); dashed lines indicate non-significant trend (p≥0.05).

The final set of sequenced 286 *K. pneumoniae* isolates represent 28.5% of the eligible biobank sampling frame (**Fig 1**). The WGS sampling fractions differed by hospital, representing 76% of all 3GCR isolates stored at Hospital A (127/167), but only 23% (63/276) from Hospital C and 17% (96/562) from Hospital B (**Table 1**). The sequenced isolates were predominantly derived from male patients (56%, 160/286) and span all age groups (see **S1 Table**). Infants (0–365 days: 81/286, 28%), adults (25–59 years: 40/286, 14%), and geriatric patients (>60 years: 47/286, 16.4%) comprised the largest age groups. Most isolates were from patients admitted to ICU (89/286, 31.1%) or medical wards (86/286, 30%), and the majority were classified as HAIs (60%, 172/286). Compared with all 3GCR isolates reported to TARSS at the three hospitals, WGS data were enriched for blood isolates (44% of WGS 3GCR isolates vs 32% of TARSS 3GCR; two-sample test, p=6.046×10^-5^), ICU patients (31% vs 19%, p=1.144×10^-5^), and younger age groups (neonates, infants and children, p=2.2×10^-16^) (**Table 1**).

### Genomic determinants underlying 3GCR

As expected, we identified an ESBL, AmpC ß-lactamase and/or carbapenemase gene in the majority of sequenced 3GCR isolates (235/286, 82.2%). For the remaining 51 isolates, we were able to retrieve inhibition zone diameters for n=43 and of these 7/43 (16.3%) had borderline measurements for ceftazidime or cefotaxime diameters (within 1–2 mm of the EUCAST 2024 breakpoints, see **Table S2**). Fourteen isolates carried porin mutations, however these mutations alone (i.e. in the absence of acquired ß-lactamase genes) are not sufficient to explain the observed resistance phenotypes. Additional unexplained AMR phenotypes were observed in 26 isolates (including 3 that had borderline assay measures), consistent with either loss of AMR plasmids associated with multiple drug classes or the presence of undetected generic mechanisms that affect multiple drugs, such as upregulated efflux (**S2 Table**). In total, this leaves 11 ESBL/AmpC/carbapenemase-free isolates where the genotype-phenotype discrepancy could not be explained by borderline phenotype measurements, plasmid loss prior to DNA extraction, or generic multidrug resistance due to efflux.

Kleborate analysis revealed substantial acquired resistance across the 286 genomes (**S1 Fig)**. Isolates carried a median of nine acquired AMR genes (Interquantile range: IQR 5–12) spanning a median of six drug classes (IQR 4–8, excluding narrow-spectrum β-lactams as this is an intrinsic or expected resistance). Determinants for aminoglycosides were most common (243/286, 85%), followed by sulfonamides (187/286, 65%), trimethoprim (171/286, 60%), narrow-spectrum β-lactams (170/286, 59%), tetracyclines (146/286, 51%), fluoroquinolones (141/286, 49%) and phenicols (131/286, 46%) (**S1 Fig**).

ESBL genes were detected in 77.0% of 3GCR isolates (**Table 2**). The most common ESBL gene detected was *bla*_CTX-M-15_ (65.4%), with other *bla*_CTX-M_ alleles accounting for the rest (**Table 2**). The distribution of these genes amongst isolates of different 3GCR profiles is shown in **Figure 3**. Most isolates carrying *bla*_CTX-M_ were resistant to both cefotaxime and ceftazidime, although some isolates carrying *bla*_CTX-M-15_ or *bla*_CTX-M-156_ were susceptible to one of these drugs.

**Table 2.** Distribution of key IJ-lactam resistance determinants among third-generation cephalosporin-resistant *K. pneumoniae* overall and stratified by hospital and specimen. Percentage and counts (n/N) indicating the fraction of isolates carrying each gene or gene family among 286 sequenced isolates, overall and stratified by hospital and specimen (blood or urine); isolates may contribute to multiple rows (i.c, co-carriage of ESBL and carbapcncmasc genes). ESBL was defined as detection of any of the following: *blac;rx.M* (any allele); *blasHV* ESBL mutation (presence ofG238S and/or E240K or substitution at codon 179 (A/T’/0/N), 169, or 148; *blaTEM* ESBL mutation (presence of any of G238S, Rl64S, El04K, E240K, A237T, or Ml82T). Carbapenemase was defined as detection of any kno, vn carbapenemase gene (those detected all belonged to either *blaNDM* or blaoXA-18,,,ikcgene families). AmpC was defined as detection of *blacMY, blaoHA,* or b/aLAP; porin mutations indicate mutations in OmpK35 and/or OmpK36. *indicate Holm-adjusted p<0.05. Hospital differences tested per gene (□^2^test or Fisher’s exact test when expected cell counts ,vere <5). For genes,vith a significant global p-value, Holm- adjusted pairwise comparisons used Fisher’s exact tests. • in hospital cells indicate differences from≥1 other hospital. No gene showed a statistically significant difference between blood and urine after Holm correction across genes. Denominators: overall= 286, Hospital A n=J27, Hospital B n=96, Hospital C n=63, Blood= 128, Urine= 158.

| Feature | Gene/ Family | Overall | Hospital |  |  | Specimen |  |
| --- | --- | --- | --- | --- | --- | --- | --- |
|  |  |  | Hospital A | Hospital B | Hospital C | Blood | Urine |
| ESBLs | Any ESBL* | 77%<br>(221/286) | 88%<br>(112/127)* | 71%<br>(68/96) | 65%<br>(41/63) | 75.7%<br>(97/128) | 78.4%<br>(124/158) |
|  | CTX-M-15* | 65.4%<br>(187/286) | 80.3%<br>(102/127)* | 50.0%<br>(48/96) | 58.7%<br>(37/63) | 64.1%<br>(82/128) | 66.5%<br>(105/158) |
|  | CTX-M-14* | 8.0%<br>(23/286) | 0.8%<br>(1/127) | 20.8%<br>(20/96)* | 3.2%<br>(2/63) | 8.6%<br>(11/128) | 7.6%<br>(12/158) |
|  | CTX-M-3 | 0.7%<br>(2/286) | 1.6%<br>(2/127) | 0.0%<br>(0/96) | 0.0%<br>(0/63) | 0.8%<br>(1/128) | 0.6%<br>(1/158) |
|  | CTX-M-156 | 0.3%<br>(1/286) | 0.8%<br>(1/127) | 0.0%<br>(0/96) | 0.0%<br>(0/63) | 0%<br>(0/128) | 0.6%<br>(1/158) |
|  | SHV mutation | 1.7%<br>(5/286) | 3.9%<br>(5/127) | 0.0%<br>(0/96) | 0.0%<br>(0/63) | 1.6%<br>(2/128) | 1.9%<br>(3/158) |
|  | TEM mutation | 1.0%<br>(3/286) | 0.8%<br>(1/127) | 0.0%<br>(0/96) | 3.2%<br>(2/63) | 0.8%<br>(1/128) | 1.3%<br>(2/158) |
| Carbapenemase | Any carbapenemase* | 19.6%<br>(56/286) | 3.9%<br>(5/127) * | 41.7%<br>(40/96)* | 17.5%<br>(11/63)* | 24.2%<br>(31/128) | 15.8%<br>(25/158) |
|  | OXA-48-like family* | 15.0%<br>(43/286) | 0.0%<br>(0/127) * | 34.4%<br>(33/96)* | 15.9%<br>(10/63)* | 17.2%<br>(22/128) | 13.3%<br>(21/158) |
|  | OXA-48* | 14.7%<br>(42/286) | 0.0%<br>(0/127) * | 33.3%<br>(32/96)* | 15.9%<br>(10/63)* | 17.2%<br>(22/128) | 12.7%<br>(20/158) |
|  | OXA-204 | 0.3%<br>(1/286) | 0.0%<br>(0/127) | 1.0%<br>(1/96) | 0.0%<br>(0/63) | 0.0%<br>(0/128) | 0.6%<br>(1/158) |
|  | NDM family* | 8.4%<br>(24/286) | 3.9%<br>(5/127) | 18.7%<br>(18/96)* | 1.6%<br>(1/63) | 11%<br>(14/128) | 6.3%<br>(10/158) |
|  | NDM-5* | 4.5%<br>(13/286) | 0.0%<br>(0/127) | 12.5%<br>(12/96)* | 1.6%<br>(1/63) | 5.5%<br>(7/128) | 3.8%<br>(6/158) |
|  | NDM-1 | 3.8%<br>(11/286) | 3.9%<br>(5/127) | 6.2%<br>(6/96) | 0.0%<br>(0/63) | 5.5%<br>(7/128) | 2.5%<br>(4/158) |
| AmpC beta-lactamase | Any AmpC* | 9%<br>(26/286) | 4.7%<br>(6/127) * | 7.3%<br>(7/96) | 20.6%<br>(13/63)* | 8.6%<br>(11/128) | 9.5%<br>(15/158) |
|  | DHA | 4.9%<br>(14/286) | 3.9%<br>(5/127) | 3.1%<br>(3/96) | 9.5%<br>(6/63) | 1.6%<br>(2/128) | 7.6%<br>(12/158) |
|  | CMY | 3.8%<br>(11/286) | 0.8%<br>(1/127) | 4.2%<br>(4/96) | 9.5%<br>(6/63) | 6.2%<br>(8/128) | 1.9%<br>(3/158) |
|  | LAP | 0.3%<br>(1/286) | 0.0%<br>(0/127) | 0% (0/96) | 1.6%<br>(1/63) | 0.8%<br>(1/128) | 0% (0/158) |
| **Porin mutation** | Porin mutation* | 25.5%<br>(73/286) | 3.1%<br>(4/127)* | 59.4%<br>(57/96)* | 19%<br>(12/63)* | 27.3%<br>(35/128) | 23.4%<br>(38/158) |
|  | With ESBL | 5.6%<br>(16/286) | 2.4%<br>(3/127) | 11.5%<br>(11/96) | 3.2%<br>(2/63) | 3.1%<br>(4/128) | 7.6%<br>(12/158) |
|  | With Carbapenemase* | 15.0%<br>(43/286) | 0.8%<br>(1/127) | 34.4%<br>(33/96)* | 14.3%<br>(9/63)* | 19.5%<br>(25/128) | 11.4%<br>(18/158) |
| **None** | No ESBL, carbapenemase, ampC or porin mutation detected | 12.9%<br>(37/286) | 11%<br>(14/127) | 10.4%<br>(10/96) | 20.6%<br>(13/63) | 14.8%<br>(19/128) | 11.3%<br>(18/158) |

**Figure 3.**
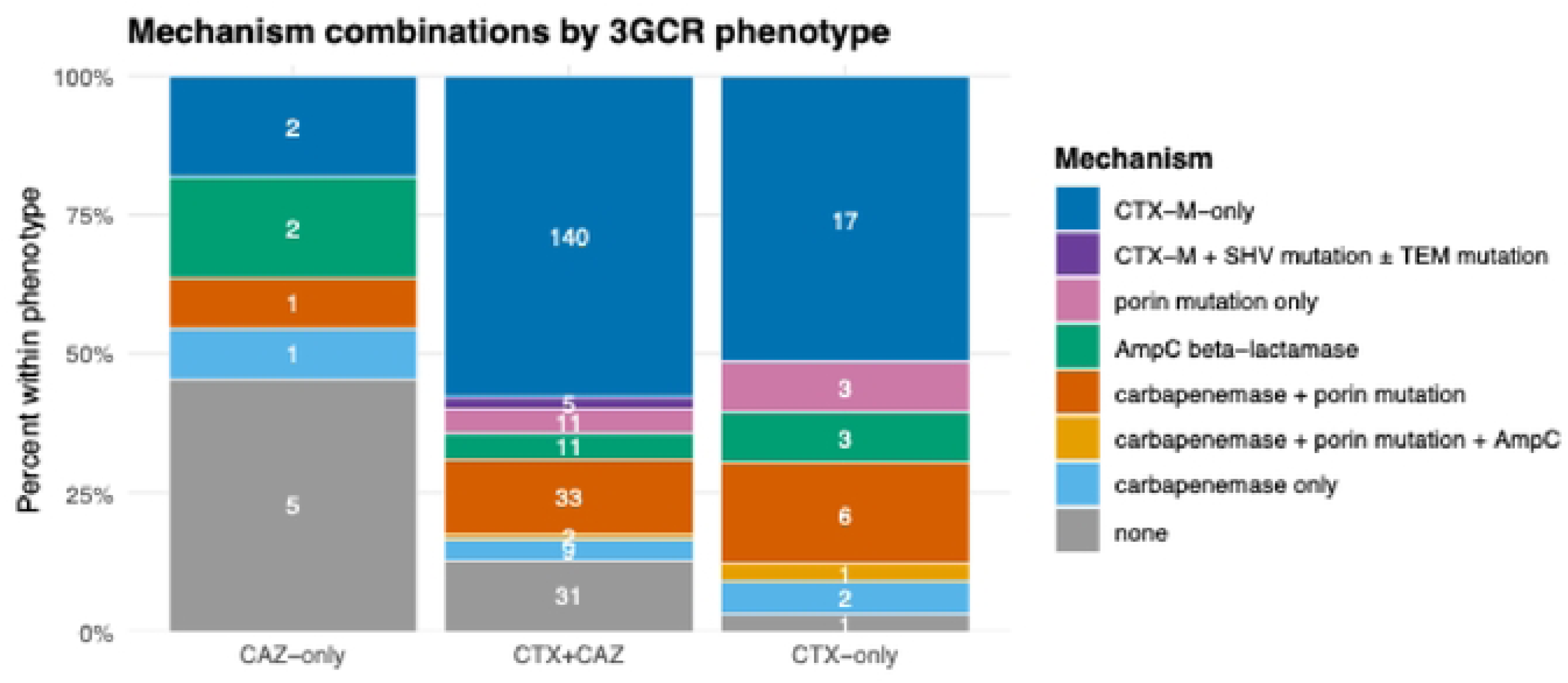
Genetic mechanisms identified among third-generation cephalosporin-resistant *K.* pneu111onu1e. Stacked bars show the distribution of known genetic mechanism identified amongst isolates of each phenotype; bars sum to 100% within each phenotype. Phenotypes were assigned from AST data (ceflazidime (CAZ) only, cefotaxime (CTX) only,or both (CTX+cAZ)). Mechanisms were categorised as defined in Table 2.

AmpC ß-lactamase genes were detected in 9% of 3GCR isolates (**Table 2**), specifically *bla*_DHA_ (4.9%), *bla*_CMY_ (3.8%) and *bla*_LAP_ (0.3%); all were resistant to both cefotaxime and ceftazidime. Carbapenemase genes were detected in 19.6% of 3GCR isolates (**Table 2)**. The most common carbapenemase genes detected were *bla*_OXA-48_ (14.7%), followed by *bla*_NDM-5_ (4.5%), and *bla*_NDM-1_ (3.8%) (**Table 2**). All *bla*_OXA-48_, *bla*_NDM-5_, and *bla*_NDM-1_ isolates were resistant to both ertapenem and imipenem. The single isolate carrying *bla*_OXA-204_ was susceptible to carbapenem agents.

There were no significant differences in the distribution of ESBL, AmpC, or carbapenemase genes between blood and urine isolates. The presence of these gene classes and some specific alleles differed significantly between hospitals (**Table 2**).

UpSet plots combining resistance determinants with cefotaxime and ceftazidime disk diffusion zones indicated that *bla*_CTX-M-15_ contributed to 3GC resistance to both drugs (**Fig. 4A–B**). For cefotaxime, *bla*_CTX-M-15_ found solo was associated with smaller disk zones (median 6 mm; IQR 6–6; positive predictive value [PPV] for resistance 0.99; n=117), and the presence of additional mechanisms, including porin alterations, AmpC, SHV/TEM mutations, did not reduce disk zones further (median 6 mm; IQR 6–6; Wilcoxon rank-sum test CTX-M-15 alone vs CTX-M-15 + other mechanisms: p=0.67). AmpC and porin alterations were individually associated with intermediate reductions in cefotaxime zones (AmpC: median 16 mm; IQR 12–16; n=13; PPV 0.85; OmpK35: median 8 mm; IQR 6–10; n=15).

**Figure 4.**
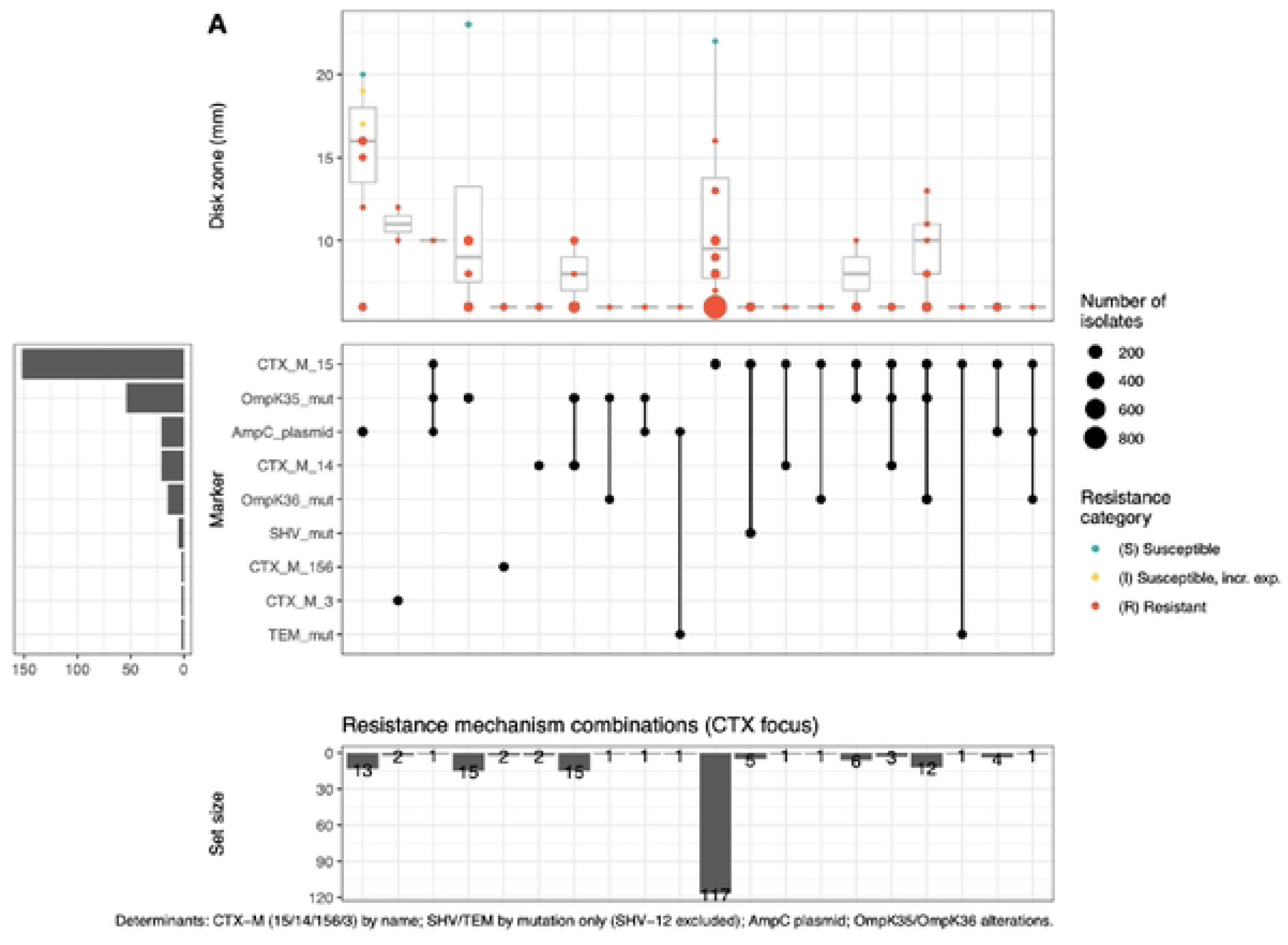
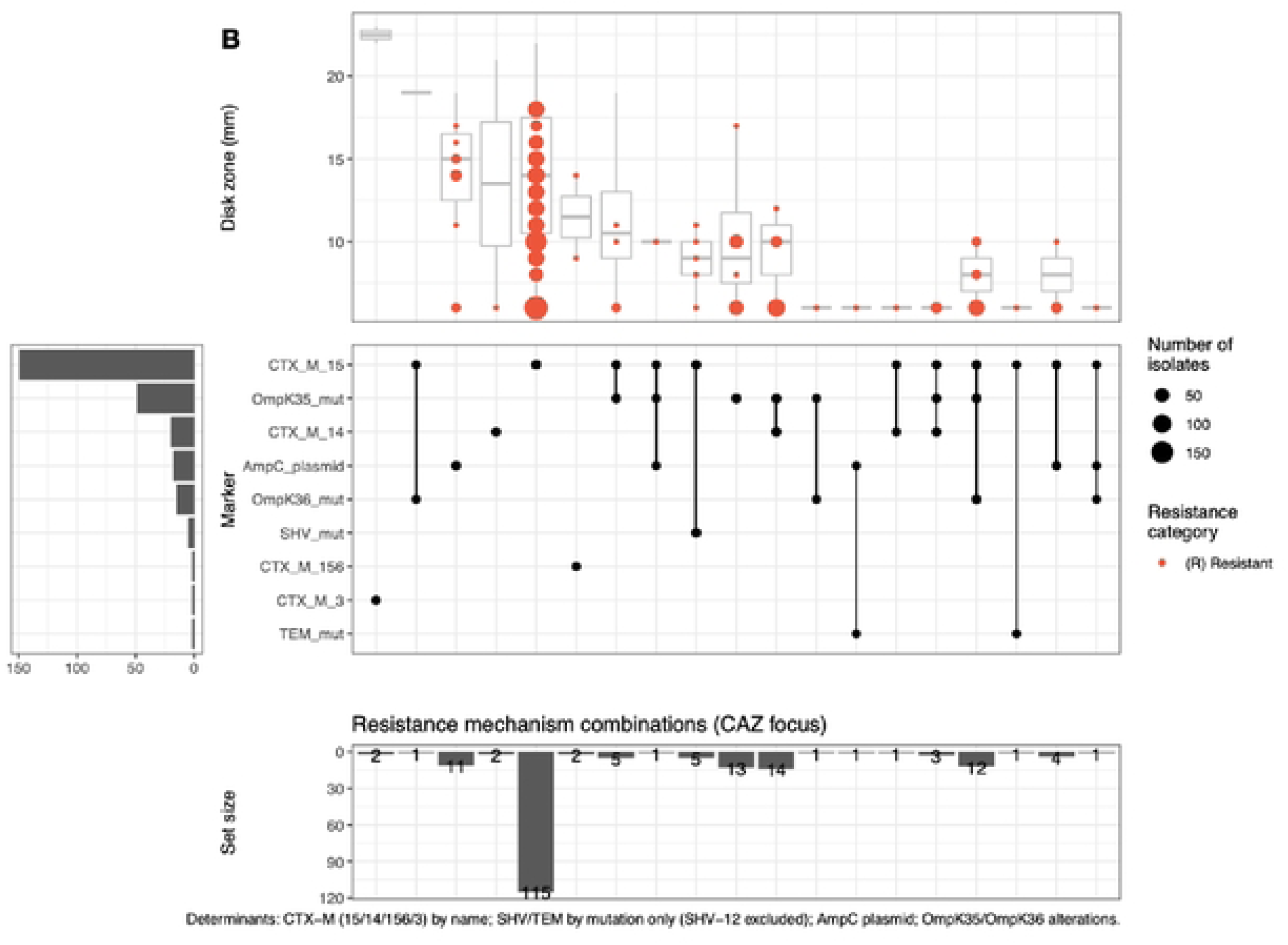
UpSet analysis of non-susceptibility versus ESBL determinants. Panels (A) Cefotaxime; (B) Ceftazidime. The top boxplot shows disk-diffusion zone diameters for the drug; isolate categories (S/1/R) interpreted using EUCAST 2024 breakpoints. Zone diameters for ceftazidime S ≥20 mm R <17mm and ccfla7.idime S:::22 mm and R<J9 (1). Vertical bars give the number of isolates with each unique combination of determinants; the dot matrix marks which determinants are present in that combination; the left bar chart shows marginal count foreach determinant. CTX-M-15, CTX-M-14, CTX-M-3, CTX-M-156; SHY and TEM by mutation only plasmid AmpC (CMY/DHA/LAP); porin alterations OmpK35 and OmpK36. Counts arc number of isolates.

For ceftazidime, *bla*_CTX-M-15_ remained the predominant mechanism (n=115) with a median disk zone of 12 mm (IQR 9–15; PPV 0.93), higher than that of cefotaxime (Wilcoxon rank-sum test p=3.6×10^-16^). Smaller disk zones were observed for isolates with ESBL genes in combination with porin disruptions, particularly OmpK35 + *bla*_CTX-M-15_ (median 6 mm, IQR 6–8; n=12; PPV 1) and OmpK35 + *bla*_CTX-M-14_ (median 6 mm; IQR 6–9; n=14; PPV 1). AmpC alone showed reduced ceftazidime zones (median 14 mm; IQR 12.5–15.5; PPV 0.9; n=11), while co-occurrence of both AmpC and *bla*_CTX-M-15_ was consistently resistant (median 6 mm; IQR 6–7; PPV 1; n= 4).

### Population structure and persistence of 3GCR *K. pneumoniae* clones

Among the 286 sequenced 3GCR *K. pneumoniae* isolates, cgMLST resolved 68 unique sublineages (SLs) (**Fig 5**). This newer lineage nomenclature (47) showed high concordance with the older 7-locus sequence type (ST) (Adjusted Rand Index [ARI] 0.887; **S4 Table**). As the neighbor-joining tree in **Fig 5** shows, most SLs (corresponding to deep branches in the tree) comprised isolates from a single hospital, however all hospitals harbored extensive diversity. SL diversity was highest at Hospital A (Simpson’s diversity 0.95; 95% CI 0.94–0.95), followed by Hospital C (0.93; 0.88–0.93), and Hospital B (0.87; 0.81–0.91) (**S5 Table**). Simpson’s diversity was equivalent and high in isolates from blood (0.96, 95% CI 0.936–0.960) or urine (0.96, 95% CI 0.938–0.958) (**S5 Table**), and no SL was significantly enriched in one specimen type after Bonferroni corrections (**S6 Table**).

**Figure 5.**
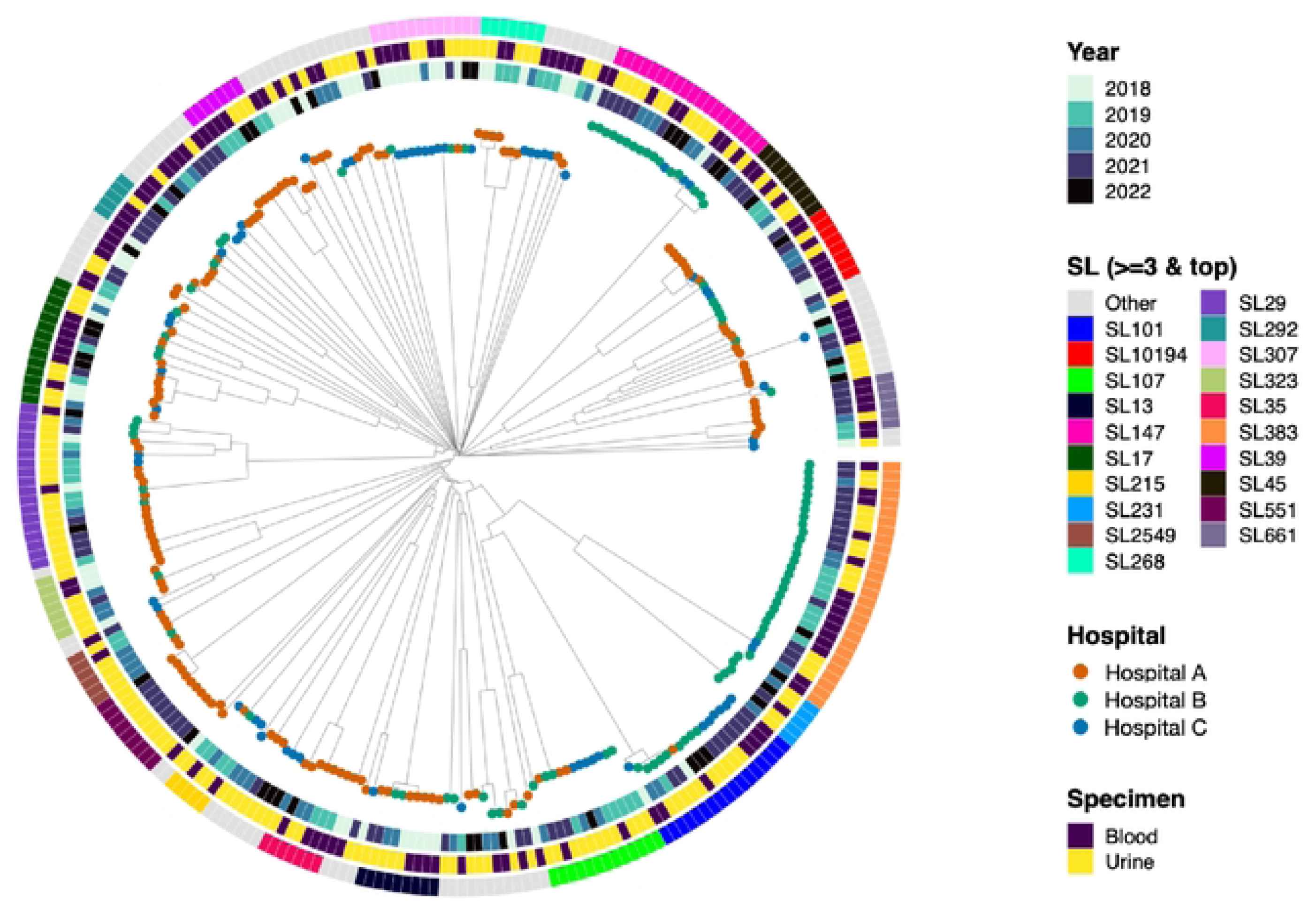
Phylogenetic tree of 3GCR *K. pneumoniae* isolates (N= 286, 2018-2022) Distance-based neighbour joining tree of 286 third-generation cephalosporin-resistant *Klebsiella pneumoniae* clinical isolates. Tips are coloured by hospital. Concentric rings from inner to outer indicate year of isolation, specimen type (blood or urine), and sublineage (SL≥ isolates and dominant lineages).

The most common ESBL gene, *bla*_CTX-M-15,_ was widely distributed across 45 unique SLs, including several that were found at multiple hospitals (e.g. SL383, SL147, SL101, SL29, SL17, SL107, SL307; see **Fig 6A**). In contrast, the rarer *bla*_CTX-M-14_ allele was confined to four SLs, concentrated in SL383 at Hospital B (where 19 SL383 isolates carried *bla*_CTX-M-14_ and 7 carried *bla*_CTX-M-15_), plus singleton isolates of SL147 (Hospital B), SL45 (Hospital A), and SL3682 (Hospital C). Notably, both SL383 and SL147 harbored *bla*_CTX-M-14_ and *bla*_CTX-M-15_, and a subset (5 SL383 and 1 SL147) carried both, suggesting parallel acquisition of distinct ESBL plasmids. The three isolates with other ESBLs each represented unique SLs, all isolated from Hospital A: SL86 and SL17 with *bla*_CTX-M-3_, and SL29 with *bla*_CTX-M-156_. ESBL-negative isolates were from diverse SLs and from all sites, and the common SLs included both ESBL-positive and ESBL-negative isolates (**Fig 6A**). For AmpC, the most common gene *bla*_DHA_ was distributed across 8 SLs in multiple hospitals, and *bla*_CMY_ appeared across 11 SLs (see **Fig 6B**). The rare *bla*_LAP_ was detected in a single isolate (SL200, at Hospital C).

**Figure 6.**
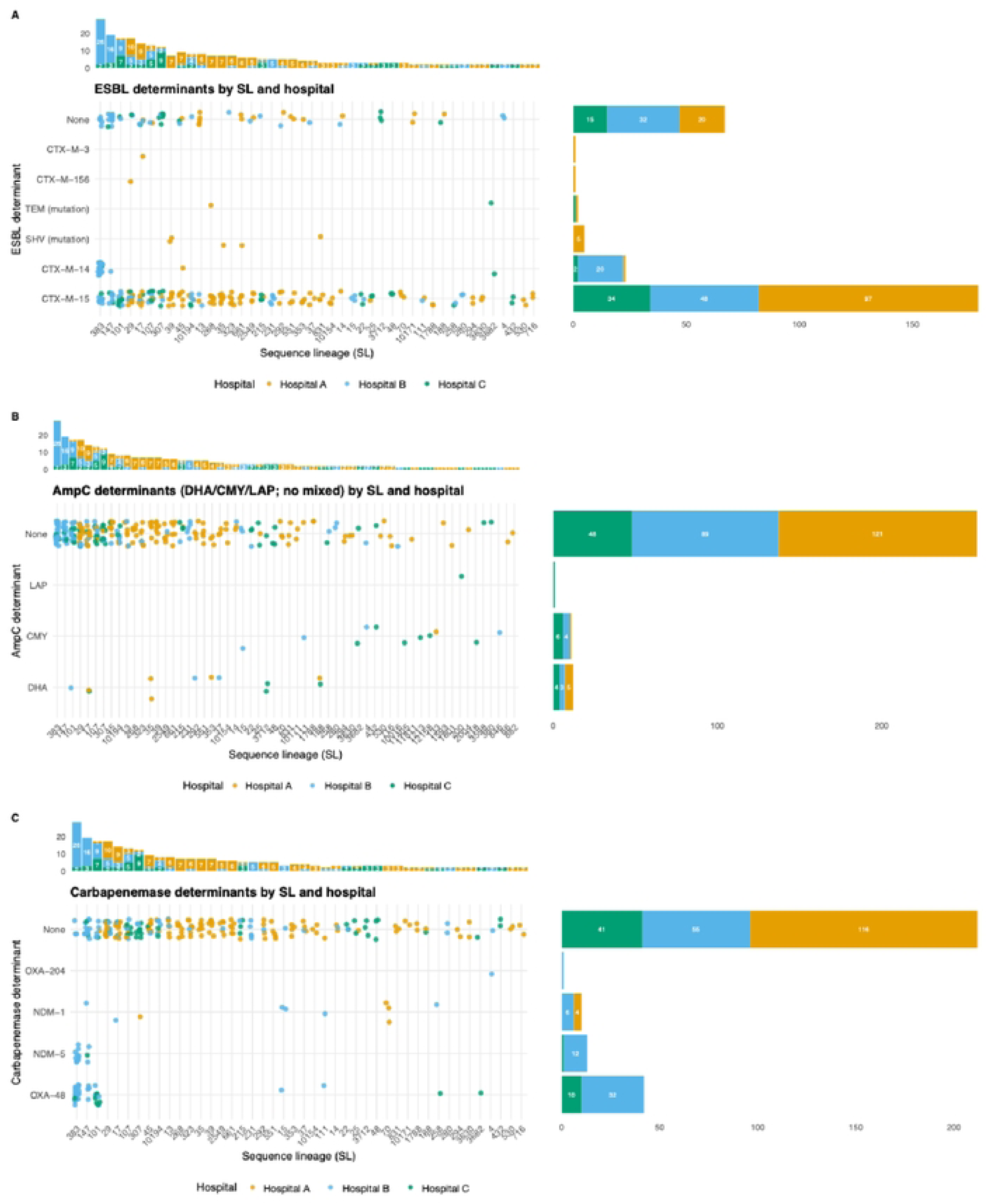
Co-occurrence of SLs, ESBL, and carbapenemase genes by sublineage and hospitals. Each point represents a ***K.*** *pneumoniae* isolate, colored by hospital and plotted by sublineage (SL; x-axis; only SLs with::: 2 isolates are shown) and acquired gene category on the y-axis. Panels show (A) ESBL (B) AmpC and (C) carbapenemase determinants. The stacked bars above each panel show the number of isolates per SL stratified by hospital. The stacked bars on the right display the number of isolates per determinant category stratified by hospital. Because isolates may carry more than one determinant, an isolate can contribute to more than one point within a panel. "None" indicates isolates with no detected gene for that category, using the following definitions: ESBL (CTX-M and SHY or TEM ESBL mutations in ESBL); AmpC (DHA, CMY, LAP); carbapenemase (OXA-48-like family and NDM variants).

Carbapenemases were detected across 12 SLs (**Fig 6C**). The most common gene, *bla*_OXA-48_ was detected in five SLs (SL383, SL101, SL111, SL147, SL15), with SL383 accounting for the largest share (n=23, detected at Hospital B (n=21) and Hospital C (n=2)). In contrast *bla*_OXA-204_ was detected in a single SL4 isolate at Hospital B. *bla*_NDM-5_ was detected in SL383 (n=10) and SL147 (n=3). Notably, both were detected examples of SL147 and SL383 isolates harboring both *bla*_OXA-48_ and *bla*_NDM-5_ and occasionally *bla*_NDM-1_, suggesting parallel acquisition of carbapenemase genes. *bla*_NDM-1_ was mainly detected at Hospital A (5/127, 3.9%) and Hospital B (6/96, 6.25%) and these were narrowly distributed across SL70 (n=3) and singleton isolates of SL307 and SL133.

Of the 68 SLs, 16 SLs were detected only once (singletons). In contrast, 13 SLs were detected at least three times but restricted to a single hospital, consistent with local persistence within sites: nine at Hospital A (SL286, SL35, SL39, and others), two at Hospital C (SL48 and SL3712), and two at Hospital B (SL231 and SL15) (**S7 Table**). Twenty-one SLs were detected in more than one hospital, and eight were present in all three (SL101, SL10194, SL107, SL17, SL215, SL29, SL307, SL45) (**Fig 7**), consistent with community circulation and repeated introductions into hospitals. SL147 was detected in Hospital B and Hospital C but was not at Hospital A (full lists in **S7 Table**).

**Figure 7.**
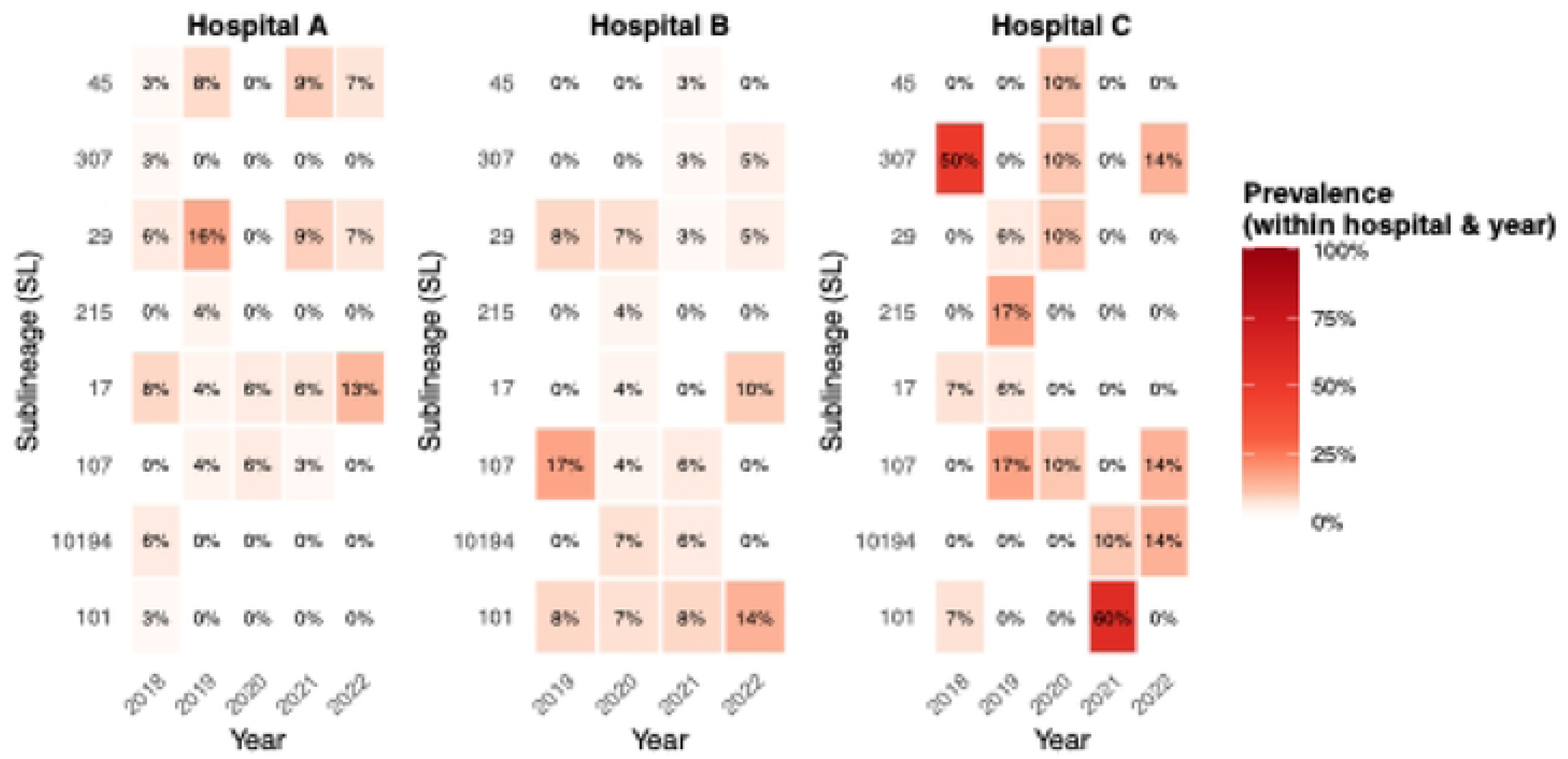
Heatmap of *K. pneunwniae* SLs detected ≥2 times in ≥2 hospitals, stratified by year and hospital. (**2018–2022**) Each panel shows the SL distribution for a different hospital. Each tile shows the number of isolates per SL and year in that hospital, coloured by prevalence of the SL amongst all isolates from Hospital (panel) and year (column). Each panel has the same rows (SL), to help compare across hospitals.

Several SLs were detected repeatedly at the same hospital over ≥3 years (**S7 Table**), suggesting sustained local persistence or recurrent reintroduction. For example, at Hospital A SL29 (2018–2022, n=10), SL17 (2018–2022, n=9), SL107 (2019–2021, n=3), and at Hospital B SL383 (2019–2022, n=26), SL147 (2020–2022, n=16), SL101 (2019–2022, n=9), and at Hospital C SL107 (2019–2022, n=5) and SL147 (2018–2022, n=3). Many of these common SLs have been reported in hospital settings in other regions, including Europe, Asia, and Africa (**S6 Fig)** (48–51).

Using our primary definition of putative transmission (HAI isolates separated by ≤20 SNPs and collected within a 56-day window), we identified 24 putative transmission clusters involving 64 isolates. These clustered isolates represent 37.2% (64/172) of sequenced 3GCR HAIs and 22.4% (64/286) of all sequenced isolates. At this cutoff, all putative transmission links were confined within hospitals, consistent with these criteria detecting within-hospital transmission rather than links resulting from transmission in the community or patient transfer between hospitals (**S10 Table**). Because sequencing covered 28.5% of overall sampling frame it is likely that the majority of HAIs occurring in the hospitals were not included in WGS and therefore were not available for consideration in the cluster analysis. It is therefore likely that many transmission links were undetected by our analysis. Accordingly, 64 clustered isolates, corresponding to 37.2% (64/172), represent a conservative minimum estimate of the fraction of 3GCR HAIs associated with nosocomial transmission. Given the low sampling fraction and lack of detailed epidemiological information or patient movement data, we cannot draw conclusions about factors contributing to transmission; however a multivariable logistic regression including bacterial (ESBL, carbapenemase, *iuc*, *ybt*) and patient factors (sex, ward, specimen, hospital) found no evidence for bacterial factors being associated with clustering; only ward and specimen type (**Fig S10**). Twenty-one percent (5/24) of putative transmission clusters included at least one carbapenemase-positive isolate, these involve SL17, SL101, SL147, and SL383 (**Table S16**).

### Serotype and virulence determinants

Capsular loci were diverse (52 loci detected) but were dominated by the KL30 locus overall (35/286, 12.2%), followed by KL64 (17/286, 6%), KL102 (16/286, 5.6%), and KL17 and KL28 (each 15/286, 5.2%) (**S17 Table**). The distribution of K loci was similar across blood and urine specimen types (**S17 Table**). K loci were largely structured by lineages: KL30 was confined to SL383, KL64 to SL147, KL17 to SL101, and KL102 to SL307 (**S12 Fig**), indicating stable capsule-lineage relationships within the population under study.

Among neonatal blood isolates (n=27), KL28 was the most frequent locus (6/27, 22.2%), detected in SL29, SL661, and SL10194, followed by KL149 (4/27, 15%) which was detected in SL39. KL122 accounted for 3/27 (11.1%), while KL110, KL16, and KL47 each accounted for 2/27 (7.4%) and were distributed across diverse SL. When compared with a recent genomic meta-analysis of neonatal invasive *K. pneumoniae* from Africa and South Asia (51), 25.9% (7/27) of neonatal blood isolates in our dataset carried K-loci included among the top-20 most common K loci in that study (KL122, KL62, KL102, and KL25).

In contrast, O-antigen diversity was limited, with eight O-types detected. O1 and O2 variants dominated: O1αß,2α (86/286, 30.1%) and O1αß,2ß (78/286, 27.3%), together accounting for 57.3% of all isolates. O2ß (43/286, 15%) and O2α (36/286, 12.6%) were also common (**S18 Table)**. The distribution of O-types was similar across specimen types (**S18 Table**). O-types were also structured by sublineage, with SL383 accounting for nearly all O1αß,2ß (27/28, 96%), SL147 for O2α (17/19, 89%), SL101 and SL307 for O2ß (16/17, 94% and 10/12, 83%, respectively), and limited O-antigen variation within lineages (**S14 Fig**).

### Virulence

Yersiniabactin (*ybt*) was the most common virulence locus detected, with similar frequencies in blood (45.3%, 58/128) and urine (51.3%, n=81/158). Aerobactin (*iuc*) was detected in 8.7% of isolates (n=25/286), with all instances belonging to the virulence plasmid KpVP-1 associated lineage, *iuc1,* and showing no association with specimen type (7.8%, n=10/128 in blood isolates vs 9.5%, n=15/158 in urine isolates). Other virulence loci were rare, with 3.1% carrying *rmpADC*, 2.7% colibactin (*clb*), and 0.7% salmochelin (*iro*). Carriage of multiple virulence determinants was rare (**S19 Table**). Eight isolates carried both *iuc* and *rmpADC* (n=3/128 blood isolates, n=5/158 urine isolates), no isolate carried the full virulence plasmid-associated complement of *iuc, iro* and *rmpADC*.

Yersiniabactin was widely distributed and detected across 36 SLs (n=139 *ybt*-positive with SL assigned). The SLs with the highest prevalence of *ybt* were SL147 (17/139, 12.2%) and SL101 (16/139, 11.5%), followed by SL17 and SL29 (each 9/139, 6.5%) and SL107 and SL45 (each 8/139, 5.8%). Among *iuc*-positive isolates (n=25), SL147 predominated (10/25, 40%), followed by SL383 (7/25, 28%) and SL101 (6/25, 24%). Two isolates belonged to additional SLs (SL17 and SL15). *rmpADC* was detected in SL383 (4/9, 44.4%), SL147 (3/9, 33.3%), SL17 (n=1), SL15 (n=1), frequently in the presence of *iuc* (8/9, 89%) (**S16–A Fig**).

As aerobactin may contribute to hyper-virulence, we examined its epidemiological distribution among our 3GCR isolates (**S20–S21 Tables**). Prevalence of *iuc* was different between hospitals (Hospital B n=16/96, 16.7%; Hospital C n=9/63, 14.3%; and Hospital A n=0/127). *Iuc*+ isolates were enriched in ICU wards (n=13/89, 14.6% vs n=12/197, 6.1% outside of ICU, p=0.0238) and were more frequent in HAI than CAI (HAI n=19/172, 11% vs CAI n=4/91, 4.4%, p=0.106). Within sublineages, CP isolates carrying *bl*a_OXA-48_ and bla_NDM-1/-5_ were identified in five SL (SL15, SL17, SL101, SL147, and SL383). SL101 (n=6), all carrying *bl*a_OXA-48_, were exclusively identified in Hospital C and ICU wards between June and October 2021. All SL383 comprised CP isolates, carrying *bl*a_OXA-48,_ were predominantly detected in Hospital B (n=6) between November 2019 and February 2021, from surgical (n=2), medical (n=3), and ICU wards (n=1). SL383 was also identified in Hospital C in November 2021 (n=1) in the ICU ward. SL147 was detected in both Hospital C (June 2022, n=1) and Hospital B (October 2022, n=1), carrying bla_OXA-48_ and bla_NDM-5_.

Two putative transmission clusters (two transmission links in total) and two singletons involved *iuc*+ isolates. The transmission clusters involved SL383, SL101, SL147, and SL17 at Hospital B and Hospital C. The hypervirulence determinants were detected in all isolates from the cluster, consistent with transmission of *iuc*+ strains.

To contextualize our local *iuc*+ SL147, which was the most common *iuc*+ lineage and involved in two clusters, we used a Mash distance-based neighbor-joining phylogeny of our SL147 isolates together with global public SL147 data. In the resulting tree (**Fig 8**), isolates from the present study were interspersed across multiple branches (red tree tips in **Fig 8**), rather than forming a single localised monophyletic clade. SL147 isolates were recovered from both clinical and non-clinical sources globally, such as animal, food, and environmental sources. Within the food sources, we identified two distant Tunisian seafood isolates collected in 2016, providing evidence of SL147 in the local Tunisian food chain (green star symbols in **Fig 8**). These isolates fall within the same clade as some of our isolates, but they do not form a tight cluster (**Fig 8**).

**Figure 8.**
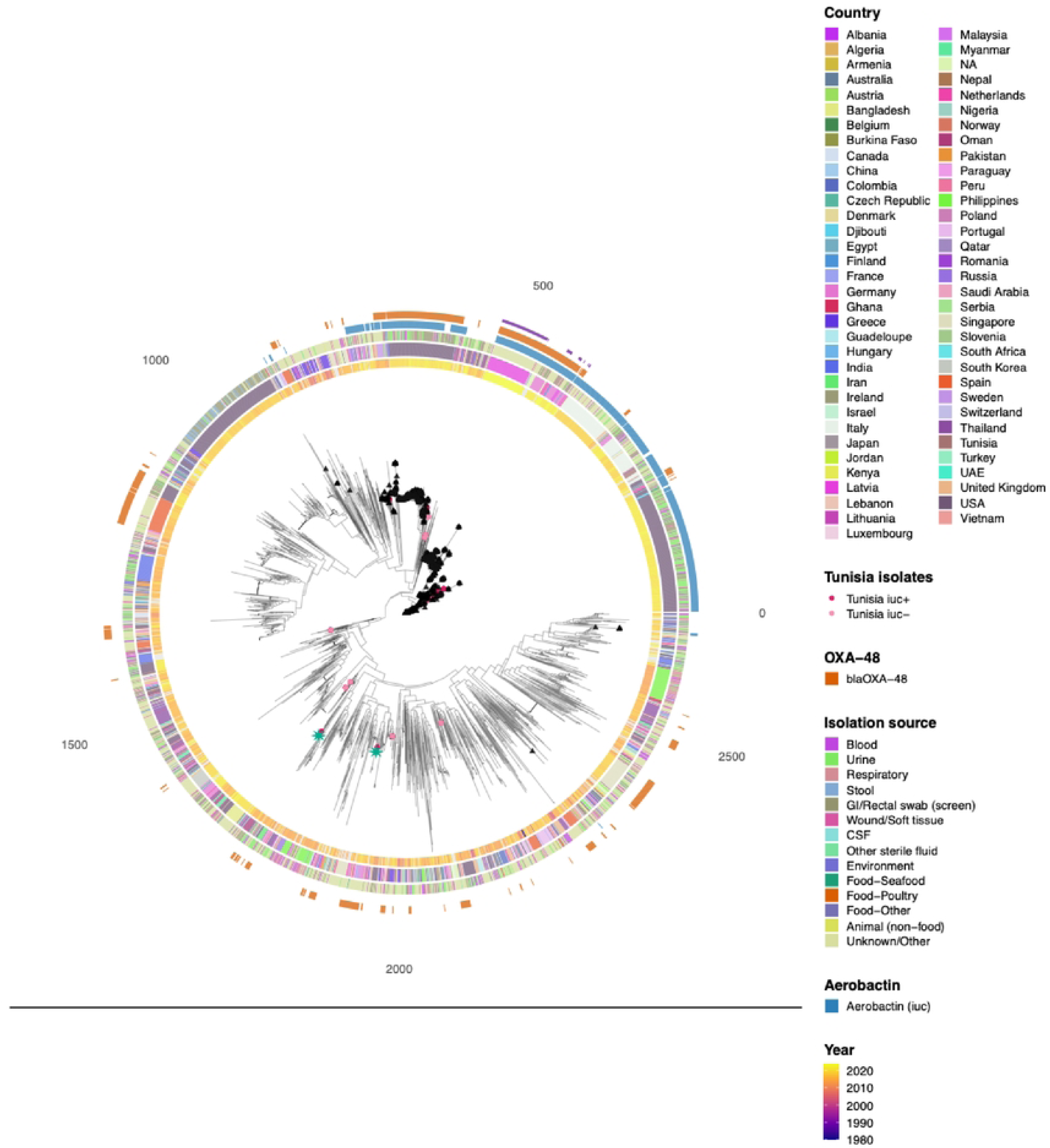
Global *K pneumoniae* ST147 phylogeny with geography and plasmid-associated traits. Neighbour-joining tree from Mash distances of global collection of ST!47 isolates (n=2,678). Rings from the tree outward show the *bla*_OXA-48_, aerobactin *(iuc)* locus, isolation source, country, and year of isolation. The triangle symbols additionally highlight non-Tunisian isolates carrying *iuc* and *blaoXA-48;* Tunisian isolates are plotted as circles (dark pink= *iuc* positive and light pink = *iuc* negative within our 3GCR landscape); the star symbol denotes Tunisian seafood samples.

Overall, 696/2678 (26%) of global SL147 genomes were *iuc*-positive, 578/2678 (21.6%) carried *bla*_OXA-48_, and 268/2678 (10%) had both. Co-occurrence of *iuc* and *bla*_OXA-48_ was identified in 51 distinct clades (hierarchical clustering height = 2.00% of maximum patristic distance), indicating that convergence is not restricted to a single lineage (blue and orange rings in **Fig 8**). The *iuc*+ isolates from Tunisia were separated into 8 distinct clades in the global tree, suggesting repeated introduction. They did not cluster with the recently reported European clade that carries a single plasmid harboring both *iuc* and *bla*_OXA-48_ (41), and we found no evidence of this plasmid in our isolates (**see Methods**).

## Discussion

Here, we analyzed whole-genome sequences for 286 confirmed 3GCR *K. pneumoniae* isolates captured by three tertiary TARSS hospitals over five years and found substantial genomic diversity within the collection. Among the confirmed 3GCR *K. pneumoniae* population, we detected high clonal diversity, with 68 sublineages spanning multiple hospitals and specimens (**Fig 5**). SL147, SL383, and SL101 were the most prevalent SLs and have been recognised in hospital outbreaks in Europe, Africa, and Asia (49,50,52). These high-risk lineages frequently harbor ESBLs, carbapenemase, *ybt*, and in some isolates, *iuc*, and *rmpADC*. The dataset represents only a subset of TARSS hospital biobank isolates and is therefore not fully representative of TARSS. Nevertheless, it provides a genomic baseline for 3GCR *K. pneumoniae* within TARSS and offers insight into circulating lineages and resistance mechanisms.

WGS confirmed 89% of the initial sample of 322 isolates were indeed *K. pneumoniae* (**Fig 1**), while the remainder were misidentified or belonged to other members of the *K. pneumoniae* Species Complex (KpSC), highlighting limitations of conventional testing within KpSC (53,54). Our 3GCR *K. pneumoniae* isolates were driven by *bla*_CTX-M-15_ ESBL (**Table 2**), as previously reported in other hospital studies (55,56). These enzymes exhibit a greater hydrolytic efficiency against cefotaxime than ceftazidime (57). OmpK35/36 are the primary entry porins for ß-lactams and their loss is likely to further reduce phenotypic inhibition zones, consistent with the small disk zones we observed in *bla*_CTX-M_ strains with porin defects in ceftazidime (**Fig 4–B**) (58). Among the 51 isolates without detectable ESBL/AmpC/carbapenemase genes, disk diffusion diameters were available for 43, 7 of which were borderline for cefotaxime or ceftazidime breakpoints (**S2 Table**). Although 14 carried porin mutations, these alone were insufficient to explain the 3GCR phenotype. Additionally, 26 isolates showed unexplained resistance across multiple antimicrobial classes, suggesting additional mechanisms not detected in our analysis. After accounting for borderline phenotypes, porin alterations, and resistance across other antimicrobial classes, only 11 isolates remained without a clear explanation for their 3GCR phenotype (**S2 Table**).

The carbapenemase gene *bla*_OXA-48_ is widely reported in North Africa and Europe and is commonly carried on pOXA-48a-like IncL/M-type conjugative plasmids, which impose low fitness costs, favoring persistence and local amplification in hospital settings (59–61). Consistent with the regional pattern, *bla*_OXA-48_ was detected among several nosocomial clusters, SL383, SL101, and SL147 (**S16 Table**), suggesting circulation of *bla*_OXA-48_ clones within the hospitals. Identifying the genetic drivers of carbapenem resistance is important for hospital stewardship programs. In settings where *bla*_OXA-48-like_ enzymes are prevalent, ceftazidime/avibactam is recommended where available (62). However, the co-occurrence of *bla*_OXA-48-like_ enzymes with *bla*_NDM_ variants in our dataset (**Fig 6C**) may compromise the activity of ceftazidime/avibactam and require alternative combination therapies (62). These findings highlight the importance of early genotypic confirmation of carbapenemase genes to guide empirical therapy and treatment decisions in hospitals.

We inferred recent transmission using thresholds of ≤20 SNPs within 56 days, after showing that a stricter SNP cut-off in our sampling frame missed plausible transmission events and larger thresholds increased cluster size via cluster-merging (**S10–S12 Tables**). Several reports have also used ≤20 SNPs to infer recent transmission within and between healthcare settings (52,63). Under this definition, 37% of HAI isolates, corresponding to 22% of all sequenced isolates, formed transmission clusters, all within hospitals (**Table 3**). This proportion is likely an underestimate, since less than a third of 3GCR isolates were sequenced. Nevertheless, Hospital B exhibited multi-ward clusters spanning ICU, medical, surgical, and pediatrics, suggesting cross-ward spread. One possible explanation for the contrast is Hospital B large hospital size (bed capacity 930) compared with other sites in this study, which may increase the opportunities for cross ward transmission (64,65). Borg *et al.* showed strong associations between intra-hospital transfers, high bed occupancy, shared-room design and increased acquisition of resistant organisms (66). In addition to hospital characteristics, lapses in IPC measures, such as hand hygiene and environmental cleaning, can amplify transmission risk (67). Multi-ward clustering at Hospital B is compatible with cross-ward spread and highlights the importance of strengthening IPC practices, including patient transfers and transfer screening, in this setting (68).

**Table 3.** Summary of within-hospital transmission clusters by site at primary SNP cut-off. Clusters were defined using an SNP ≤20 threshold and a 56-day temporal window and restricted to HAI only. *"N’* is the sequenced dataset after filtering to HAI only. "No. clusters", indicates the number of transmission clusters with ≥2 HAI isolates (HAI singletons excluded). "Isolates in clusters", is the number of HAI isolates belonging to these clusters."% isolates in clusters" is the proportion of clustered HAI isolates in clusters. "Attributable" is the proportion of HAI isolates estimated to be "secondary cases" under a one-index-case-per-cluster assumption. "Median span" and "interquartile (IQR) range" of cluster duration in days. "% Mono-ward" represents the proportion of clusters confined to a single ward (each cluster counts once).

| No. of HAI isolates sequenced (N) | No. clusters | Isolates in clusters<br>n (%) | Isolates attributable to transmission<br>n (%) | Median span (days) | IQR span (days) | Mono-ward clusters<br>n (%) |
| --- | --- | --- | --- | --- | --- | --- |
| Hospital A (N=60) | 11 | 27/60 (45%) | 16/60 (27%) | 13 | 26.5 | 8/11 (72.7%) |
| Hospital B (N =63) | 7 | 22/63 (35%) | 15/63 (24%) | 7 | 40 | 1/7 (14.3%) |
| Hospital C (N =49) | 6 | 15/49 (31%) | 9/49 (18.5%) | 9 | 13.75 | 6/6 (100%) |
| Overall (N =172) | 24 | 64/172 (37.2%) | 40/172 (23.2%) | 11 | 27 | 15/24 (62.5%) |

The proportion of convergent AMR-hypervirulent *K. pneumoniae* (3GCR + *iuc*) in our dataset (8.7%) was comparable to other regional and global estimates (7% to 12.6%) (69–72). Nonetheless, convergence is a growing threat to healthcare in Tunisia, particularly in high-risk wards, such as the ICU, due to their limited therapeutic options and their linkage to invasive diseases and high mortality (73) Since plasmids move across lineages, they can seed new convergent combinations in networks with substantial patient movement and device exposure, underscoring the necessity of their early detection and containment (74).

Capsular polysaccharide has gained traction as a therapeutic target due to its immunomodulatory characteristics and the ability of capsule-targeting antibodies to kill *K. pneumoniae* (75). The diversity and broad distribution of K and O types challenge the future of vaccine efficacy, and Tunisia is no exception. Comparing our neonatal K loci to the top repertoires reported by Stanton *et al.* in Africa and South Asia, we observed partial overlap (25%), suggesting that the antigen-targeted vaccines under development could offer partial coverage in this setting (51). Therefore, Tunisia should continue local surveillance to assess vaccine coverage and guide future development.

SL147 is a high-risk clone implicated in several hospital outbreaks worldwide (36,76). In our dataset, SL147 genomes were dispersed across the global phylogeny, consistent with multiple introductions followed by local dissemination (**Fig 8**). These isolates frequently carried plasmids such as IncHI1B (pNDM-MAR) and Col-like plasmids encoding *bla*_CTX-M-15_ and *bla*_NDM-1_ and have recently been reported to carry *bla*_OXA-48_ (77). In 2025, European surveillance reported a convergent SL147 lineage harboring an IncHI1B mosaic plasmid, aerobactin and *bla*_OXA-48_, and was responsible for cross-border spread and hospital outbreaks (41). We found no evidence of the exact European epidemic plasmid but found a related IncHI1B-family variant circulating in Tunisian hospitals. Given SL147 international movement, its propensity for acquiring additional resistance and virulence, and its presence in clinical and non-clinical reservoirs, warrants genomic surveillance within a One Health framework in Tunisia (78).

In summary, we have shown that 3GCR *K. pneumoniae* in the three TARSS hospitals dominated by *bla*_CTX-M-15_ and high-risk lineages consistent with nosocomial transmission routes. Although our SL147 isolates were not part of the cross-border European mosaic plasmid, their resemblance and mobility argue for vigilance across human, animal, food, and environmental sectors. We demonstrated that integrating WGS into TARSS is feasible in order to identify high-risk clones early in the course of an incipient epidemic. We also highlight the utility of WGS to guide stewardship and IPC policies in TARSS.

### Limitations

While this study provides unprecedented genomic resolution for 3GCR *K. pneumoniae* within Tunisian hospitals, several limitations should be acknowledged. First, the study was retrospective; we only sequenced archived clinical isolates, and some were unavailable due to biobanking losses. In 2018, Hospital B was unable to retrieve any isolates, and the Hospital C metadata was incomplete for age. Therefore, the cohort is not population-representative of the TARSS sentinel hospitals and may overrepresent certain age groups, wards, and specimen types. Second, our genomic and transmission analyses are limited to 3GCR *K. pneumoniae* with incomplete sequencing and will miss links; therefore, these transmission estimates should not be extrapolated to all *K. pneumoniae* infections in the hospitals. Third, patient-level metadata, including movement between wards, antibiotic exposure, device use timelines, and contact tracing, were unavailable. As a result, our cluster alignments rely on core-genomic proximity within time windows and coarse ward and collection metadata. This limited our ability to infer transmission pathways or directionality. Finally, sequencing was short-read only, reflecting current capacity at the NRL. Illumina short read sequencing constrains plasmid reconstruction and resolution of repetitive or mobile elements, and may fragment loci across contigs.

## Conclusion

To our knowledge, this is the first WGS analysis undertaken within the Tunisian hospital-based national AMR surveillance program. We demonstrate that the burden of 3GCR *K. pneumoniae* in participating hospitals is driven by a polyclonal population concentrated for internationally recognized high-risk clones (notably SL147, SL383, and SL101), and the dominant carbapenemase detected was *bla*_OXA-48_. Aerobactin-positive 3GCR and carbapenemase-producing strains were present but were interspersed in the phylogeny suggesting they are not yet locally established as intensively transmitting clones. Globally, our Tunisian SL147 isolates were widely distributed and harbored IncHI1B-family variants rather than the exact mosaic backbone reported in Europe.

Integrating WGS within TARSS has the potential to add resolution to transmission, support early detection of genotypes to guide empirical treatment, and longitudinal tracking of lineages, plasmids, and convergence. Delivering this at a national scale, through a representative multiregional sampling frame, will strengthen IPC and antimicrobial stewardship and support progress towards NAP of AMR objectives.

## Data Availability

The WGS sequences generated in this study are publicly available in the European Nucleotide Archive (ENA) under BioProject PRJEB98550. Individual sample accession numbers are provided in the Supplementary Dataset along with de-identified metadata supporting the findings of this study. The analysis code used for data processing, statistical analysis, and visualization, are available in a public repository at: https://zenodo.org/doi/10.5281/zenodo.19055814 To minimise the risk of participant re-identification, certain patient-level variables are not publicly shared (age, ward, and date of admission). Access to additional metadata required for reproducibility may be requested from the relevant institutional authority, subject to ethical approval.

https://www.ebi.ac.uk/ena/browser/view/PRJEB98550

## Acknowledgements

We thank our collaborators from the Tunisian hospitals and the NRL for their invaluable contributions, especially for providing the data. This work was conducted as part of TARSS under the supervision of the NRL. We further acknowledge the sustained support of the NRL and the MOH in Tunisia for their leadership and continued commitment to AMR surveillance activities. We also extend our appreciation to our collaborators at Quadram Institute Bioscience for performing the library preparation and sequencing.

## Author contributions

Conceptualization, DI, KEH; methodology, DI, KEH, EFN; data provision, IB, MZ, HS, LT; epidemiological and microbiological data collection from the participating institutions, SD, LK, KM; formal analysis, DI; sequencing, DJB, CJM; visualization, DI; writing—original draft preparation, DI; writing—review and editing, DI, KEH, EFN, LTP, IB, HS, MZ, LT, WA, SD, LK, KM, DJB, CJM; supervision, KEH. All authors have read and approved the final version of the manuscript.

## Conflict of interest

The authors declare no conflict of interest.

## Funding

The author(s) received no specific funding for this work.

## Supporting information

**S1 Table. Comparison of TARSS surveillance isolates and sequenced (WGS) isolates by hospital and clinical variables, 2018–2022**

**S2 Table. Genotype-phenotype concordance for co-resistance determinants amongst 51 third-generation cephalosporin-resistant *K. pneumoniae* isolates lacking acquired ESBL, AmpC, or carbapenemase genes, Tunisia, 2018–2022**

**S3 Table. Sublineages (SL) heterogeneity by Sequence Type (ST)**

**S4 Table. Agreement between Sequence type and SL labels (Adjusted Rand Index) and number of isolates evaluable for nearest-neighbour analysis.**

**S5 Table. Diversity of *K. pneumoniae* sublineages (SL) within each hospital site and specimen type, measured by Simpson’s index (95% CI).**

**S6 Table. Sublineages associated with Specimen type (urine and blood) among third-generation cephalosporin-resistant *K. pneumoniae***

**S7 Table. Sublineages (SL) occurrence by hospital and years of detection, 2018-2022**

**S8 Table. Sublineages (SL) detected in both blood and urine among 3GCR K. pneumoniae, 2018-2022**

**S9 Table. Specimen-specific persistence of SLs among 3GCR *K. pneumoniae*, 2018-2022**

**S10 Table. Transmission clustering metrics by SNP threshold and temporal window (28,56, and 84 days)**

**S11 Table. Window-stability of transmission metrics across SNP thresholds (8 vs 12 weeks)**

**S12 Table. Stability of cluster membership between adjacent SNP thresholds (56-day window)**

**S 13 Table Jaccard similarity of lineage composition across adjacent SNP thresholds (56-day window)**

**S14 Table. Coverage of lineage set at each SNP threshold relative to the final (40-SNP) set, at 56 days**

**S15 Table. Observed same-ward concordance vs permutation null under candidate cut-offs (≤15 or ≤20 SNP; ≤56 days)**

**S16 Table. Transmission clusters (SNP ≤20 within 56 days) among 3GCR *K. pneumoniae* (2018–2022)**

**S17 Table. K-loci distribution among 3GCR *K. pneumoniae* isolates (N = 286) S18 Table. O-type distribution among 3GCR *K. pneumoniae* isolates (N = 286)**

**S19 Table Virulence locus prevalence (single loci and combinations) overall and by specimen**

**S20 Table. Characteristics of convergent (iuc-positive) *K. pneumoniae* isolates**

**S21 Table. Distribution of convergent isolates across SL, hospital, specimen, ward, patient type, and year**

**S22 Table. Convergent isolates within SNP-defined transmission clusters (≤20 SNP, ≤56 days; HAI only)**

**S23 Table. Bandage results for all SL147 isolates**

**S1 Fig. Frequency of isolates carrying any acquired determinant by Kleborate class**

**S2 Fig. Temporal trends in ESBL categories by hospital and specimen**

**S3 Fig. Temporal trends of AmpC genes by hospital and specimen**

**S4 Fig. Temporal trends of carbapenemase genes by hospital and specimen**

**S5 Fig. Detection of Tunisian sublineages (SL) across global genomic datasets**

**S6 Fig. Heatmap of *K. pneumoniae* SLs detected ≥3 times in any specimen, stratified by year and specimen (2018–2022)**

**S7 Fig. Impact of SNP cut-off and temporal window on transmission clustering: yield, count, and duration**

**S8 Fig. Ward localisation and duration of healthcare-associated transmission clusters of 3GCR *K. pneumoniae* by hospital (SNP≤20; 56 days; HAI only)**

**S9 Fig. Predictors of transmission-cluster membership (≤20 SNP, ≤56 days)**

**S10 Fig. AMR and virulence composition of transmission clusters by sublineage and hospital (SNP ≤ 20; ≤ 56 days; HAI only)**

**S11 Fig. Frequency of K-locus among blood and urine 3GCR *K. pneumoniae* isolates with sublineage composition**

**S12 Fig. Frequency of K-loci across age groups stratified by sublineage (SL)**

**S13 Fig. Frequency of O-type among blood and urine 3GCR *K. pneumoniae* isolates with sublineage composition**

**S14 Fig. Frequency of O-type across age groups stratified by sublineage (SL)**

**S15 Fig. Virulence loci across sublineages**

**S16 Fig. Bandage assembly graph for (SL147) isolate number 1481: IncHI1B(pNDM-MAR), blaOXA-48 and aerobactin and Oligo detected; loci on separate components**

## Notes

### Competing Interest Statement

The authors have declared no competing interest.

### Author Declarations

Ethical approval for this study was obtained from the Ministry of Health in Tunisia, the Ethics Committee of Charles Nicolle Hospital, Tunis, and the London School of Hygiene & Tropical Medicine (LSHTM) Observational/Interventions Research Ethics Committee. The study used retrospective, de-identified surveillance data and archived bacterial isolates. A waiver of individual informed consent was granted by the relevant ethics committees.

